# High-resolution African HLA resource uncovers *HLA-DRB1* expression effects underlying vaccine response

**DOI:** 10.1101/2022.11.24.22282715

**Authors:** Alexander J. Mentzer, Alexander T. Dilthey, Martin Pollard, Deepti Gurdasani, Emre Karakoc, Tommy Carstensen, Allan Muhwezi, Clare Cutland, Amidou Diarra, Ricardo da Silva Antunes, Sinu Paul, Gaby Smits, Susan Wareing, HwaRan Kim, Cristina Pomilla, Amanda Y. Chong, Debora Y.C. Brandt, Rasmus Nielsen, Samuel Neaves, Nicolas Timpson, Austin Crinklaw, Cecilia S. Lindestam Arlehamn, Anna Rautanen, Dennison Kizito, Tom Parks, Kathryn Auckland, Kate E. Elliott, Tara Mills, Katie Ewer, Nick Edwards, Segun Fatumo, Sarah Peacock, Katie Jeffery, Fiona R.M. van der Klis, Pontiano Kaleebu, Pandurangan Vijayanand, Bjorn Peters, Alessandro Sette, Nezih Cereb, Sodiomon Sirima, Shabir A. Madhi, Alison M. Elliott, Gil McVean, Adrian V.S. Hill, Manjinder S. Sandhu

**Affiliations:** Wellcome Centre for Human Genetics, University of Oxford, UK; Big Data Institute, Li Ka Shing Centre for Health Information and Discovery, University of Oxford, Oxford, UK; Institute of Medical Microbiology and Hospital Hygiene, University Hospital of Düsseldorf, Heinrich Heine University Düsseldorf; Genome Informatics Section, Computational and Statistical Genomics Branch, National Human Genome Research Institute, Bethesda, Maryland, USA; Wellcome Sanger Institute, Hinxton, Cambridge, UK; Medical Research Council/Uganda Virus Research Institute and London School of Hygiene & Tropical Medicine Uganda Research Unit, Entebbe, Uganda; South African Medical Research Council Vaccines and Infectious Diseases Analytics Research Unit, University of the Witwatersrand, Johannesburg, South Africa; Groupe de Recherche Action en Santé (GRAS) 06 BP 10248 Ouagadougou, Burkina Faso; Division of Vaccine Discovery, La Jolla Institute for Immunology, La Jolla, California, USA; National Institute for Public Health and the Environment, Bilthoven, The Netherlands; Microbiology Department, John Radcliffe Hospital, Oxford University NHS Foundation Trust, Oxford, UK; Histogenetics, New York, USA; Department of Integrative Biology, University of California at Berkeley, California, USA; Avon Longitudinal Study of Parents and Children at University of Bristol, Bristol, UK; Population Health Sciences, Bristol Medical School, University of Bristol, Bristol, UK; MRC Integrative Epidemiology Unit, University of Bristol, Bristol, UK; Department of Infectious Disease, Imperial College London, UK; The Jenner Institute, University of Oxford, UK; The Department of Non-communicable Disease Epidemiology, London School of Hygiene and Tropical Medicine London, London, UK; Tissue Typing Laboratory, Cambridge University Hospitals NHS Foundation Trust, Cambridge, UK; University of California San Diego, La Jolla, CA, USA; Department of Clinical Research, London School of Hygiene & Tropical Medicine, London, UK; Department of Epidemiology & Biostatistics, School of Public Health, Imperial College London, UK

## Abstract

How human genetic variation contributes to vaccine immunogenicity and effectiveness is unclear, particularly in infants from Africa. We undertook genome-wide association analyses of eight vaccine antibody responses in 2,499 infants from three African countries and identified significant associations across the human leukocyte antigen (HLA) locus for five antigens spanning pertussis, diphtheria and hepatitis B vaccines. Using high-resolution HLA typing in 1,706 individuals from 11 African populations we constructed a continental imputation resource to fine-map signals of association across the class II HLA observing genetic variation explaining up to 10% of the observed variance in antibody responses. Using follicular helper T-cell assays, *in silico* binding, and immune cell eQTL datasets we find evidence of *HLA-DRB1* expression correlating with serological response and inferred protection from pertussis following vaccination. This work improves our understanding of molecular mechanisms underlying HLA associations that should support vaccine design and development across Africa with wider global relevance.

**Teaser:** High-resolution typing of HLA diversity provides mechanistic insights into differential potency and inferred effectiveness of vaccines across Africa.

## Introduction

Vaccination is one of the most cost-effective methods for preventing disease caused by infections world-wide^1^. The strategy has been successful for eradicating smallpox, and also reducing morbidity and mortality associated with other infections, many of which were commonplace in the pre-vaccination era^2^. Such diseases include diphtheria (a toxin-mediated disease caused by *Corynebacterium diphtheriae*), pertussis (another toxin-mediated disease caused by *Bordetella pertussis*) and measles, all of which have vaccines delivered in infancy as part of the expanded programme on immunisation (EPI).

Despite the unquestionable success of vaccination, significant challenges remain both for maintaining control of vaccine-preventable diseases, and in the development of vaccines against other diseases that are more challenging to target in successful vaccination strategies. For example, epidemics of pertussis are being increasingly reported in vaccinated communities^3^. The incidence of these vaccine failures appears to have increased since the move away from whole-cell, to acellular (multi-antigen) pertussis preparations, a decision largely made on the basis of increased reactogenicity following whole-cell vaccination^4^. However, the specific mechanisms underlying the increase in rates of failures remain unclear, and several countries (particularly in Africa) continue to use whole-cell preparations. Furthermore, it is well recognised that several infectious diseases pose particular problems for vaccine development including tuberculosis^5^, malaria^6^, human immunodeficiency virus^7^, and even SARS-CoV-2 where increasing reports of vaccine breakthrough infection are being reported as early as six months following two doses of vaccine^8^. Amongst the multitude of challenges posed in these diverse development efforts, two distinct challenges are common amongst both the vaccine-preventable and more challenging diseases. Firstly, the antigens to target and the ideal components of the immune response to stimulate to induce protection – so called correlates of protection – are often difficult to define^9^. Secondly, given the necessary world-wide scope of delivery required for many vaccines and the diversity of factors that may influence immune response to vaccination, understanding population differences in risks of vaccine failure is important, particularly in low-to-middle income countries where reporting of failures may not be effectively captured, and where the burden of vaccine preventable diseases is frequently the highest.

One feature of population differences that has been under-studied to date is human genetic variation. It has been recognised for decades that variation across the major histocompatibility complex (MHC), known in humans as the human leukocyte antigen (HLA) locus, is associated with differential response and failure to respond to the hepatitis B surface antigen (HBsAg) vaccine^10^, as well as responses against tetanus toxin (TT)^11^ and measles vaccines (MV)^12^. These findings are in keeping with the well-known association of the locus with susceptibility to multiple other infectious and autoimmune diseases^13–15^. We have recently found evidence that carriage of specific *HLA* gene product alleles (HLA-DQB1*06 in particular) may improve SARS-CoV-2 vaccine immunogenicity and reduce the risk of breakthrough infection with COVID-19 post-vaccination^16^. Despite the recognition of these associations, it has not been possible to elucidate the precise underlying causal mechanisms. The presence of *HLA* genes across this locus leads to the speculation that differential peptide binding is responsible. However, the high concentration of genes in the region, the high levels of genetic diversity and epistatic interactions among *HLA* loci within long stretches of linkage disequilibrium pose substantial challenges to fine-mapping any association signals reliably. Any mapping and downstream mechanistic interpretation is particularly challenging in populations hitherto under-represented in global genetic studies. Despite statistical and computational advances for HLA biology using methods such as HLA imputation applied to common autoimmune diseases including multiple sclerosis^17^ and inflammatory bowel disease^18^ and a limited number of infectious agents such as HIV-1^19^, progress has largely been restricted to populations of European ancestry. Given the worldwide, standardised delivery of vaccines, studying vaccine response heterogeneity in African populations offers the opportunity to not only understand the influence of host genetics in this diverse, infection burdened and vulnerable set of populations, but also to improve our understanding of mechanisms of vaccine response and thus open avenues for vaccine development for other infectious diseases of importance.

Here we present our findings from a set of genome-wide association studies of diverse vaccine responses in African infants. We find associations across the HLA with five of eight measured antigens delivered as part of the EPI programme. In order to understand the implications and mechanisms underlying these associations we developed a comprehensive high-resolution HLA reference panel for imputation and a suite of expression quantitative trait loci (eQTL) resources for HLA. Alongside of peptide binding and immunological assays we highlight *HLA-DRB1* expression as a possible factor associated with differential inferred protection against pertussis as well as antibody responses against both pertussis and diphtheria antigens. This study highlights the importance of accounting for genetic diversity in vaccine design, deployment and universal effectiveness and provides a framework to support optimal population-adjusted vaccine design and development across Africa and worldwide.

## Results

### HLA associations with diverse vaccine responses in African infants

Given limited understanding of the contribution of host genetics to variation in response and effectiveness of the most widely delivered vaccines in the world, and the need to understand such responses in under-represented populations of the world, we tested for association between vaccine antigen responses and genetic variants (17 million variants typed and imputed with the merged 1000 Genomes – 1000Gp3 – and African Genome Diversity Project – AGDP – reference panel^20^) in 2,499 infants recruited from three African countries (Burkina Faso (BF), South Africa (SA) and Uganda (UG) defined as the *VaccGene* cohorts, **Fig. 1A**). The vaccine responses included were immunoglobulin G (IgG) antibody levels against eight vaccine antigens (diphtheria toxin (DT); pertussis toxin (PT), filamentous haemagglutinin (FHA), and pertactin (PRN); tetanus toxin (TT); *Haemophilus influenzae* type b (Hib); measles virus (MV); and hepatitis B surface antigen (HBsAg)). The demographics of the *VaccGene* populations are described in **Table S1** and a summary of the participating individuals and stringent quality control is provided in **Fig. S1A**, Methods and **Tables S2** and **S3**. The IgG traits were normalised (using inverse normal transformation, with distributions represented in **Fig. S1B)** and association testing was performed with time between last vaccine and blood sample included as a fixed effect covariate which was shown to be inversely correlated with all traits with response to DT as an exemplar in **Fig. S1C**. A genetic relatedness matrix was included in the association model as a random effect covariate using a pooled linear mixed model^21^. We identified significant evidence of association within the HLA region for five vaccine responses including pertussis toxin (PT), pertussis filamentous haemagglutinin (FHA), pertussis pertactin (PRN), diphtheria toxin (DT) and HBsAg (**Fig. 1B** and **Additional Data Table 1**). The patterns of pooled association statistics were different across each of the tested traits but all index variants with the smallest *P*-value were centred on the class II HLA region and particularly the *HLA-DRB1* (rs73727916 for PT, beta=0.33, *P*=1.9×10^-27^; rs34951355 for DT, beta=-0.56, *P*=1.5×10^-26^; rs6914950 for HBsAg, beta=0.35, *P*=9.0×10^-13^) and *HLA-DQ* (rs1471103672 for FHA, beta=-0.30, *P*=9.8×10^-16^; rs147857322 for PRN, beta=0.37, *P*=4.2×10^-23^) gene loci. No associations were observed outside of the HLA either at an individual or pooled cohort level for any trait and no associations were observed across the genome for MV or TT responses (**Figs. S1D** and **S1E**). This is the first report to our knowledge that demonstrates the importance of genetic variation in influencing the response to vaccine antigens in African infants.

**Fig. 1.**
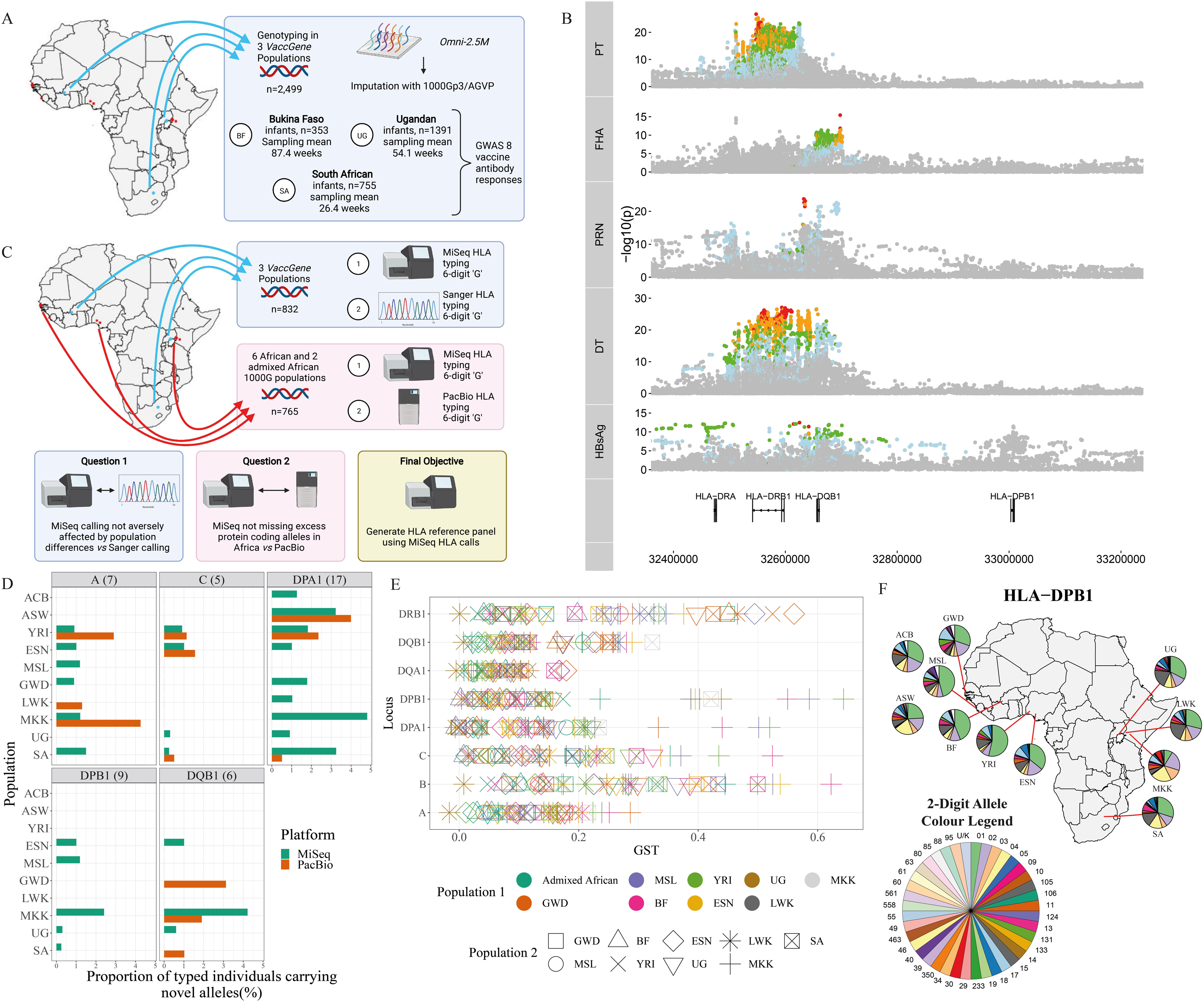
HLA associations with diverse vaccine responses in African infants and the diversity of HLA alleles across Africa. **(A)** A schematic of the experimental design for the VaccGene project genotyping DNA from 2,499 infants across three African sites and testing for association with eight vaccine antibody responses. **(B)** A regional association plot of pooled genetic association statistics of imputed and directly genotyped variants tested for association with five vaccine antigen responses demonstrating unique patterns of association across the class II HLA region. Points are coloured by linkage disequilibrium (r^2^) with the index variant in each analysis across all three populations: red (0.8-1), orange (0.6-0.8), green (0.4-0.6), blue (0.2-0.4) and grey (<0.2). **(C)** Schematic of experimental design to call HLA allelic diversity using DNA from 1,597 individuals across nine sites in Africa and two admixed African-American populations. **(D)** The proportion of individuals in each population with novel alleles confidently called using either MiSeq or PacBio calling pipelines. Total numbers of typed individuals can be found in **Table S5**. **(E)** Measures of differentiation between African populations for eight *HLA* genes across class I and II loci. Estimates, in G_ST_, are between pairs of populations with the first population represented as the colour and the second as a shape allowing a determination of the combination of populations through colour and shape. **(F)** Pattern of differentiation of HLA-DPB1 2-digit alleles with frequencies plotted as pie-charts by population across Africa. ACB: African Caribbean in Barbados; ASW: African Ancestry in Southwest USA; BF: Burkina Faso; ESN: Esan in Nigeria; GWD: Gambian in Western Division, The Gambia – Mandinka; LWK: Luhya in Webuye, Kenya; MKK: Maasai in Kinyawa, Kenya; MSL: Mende in Sierra Leone; SA: South Africa; UG: Uganda; YRI: Yoruba in Ibadan, Nigeria.

### High resolution HLA typing across Africa

In order to move towards an increased understanding of the mechanisms underlying the HLA associations observed with the vaccine antigens we first sought to determine the relationship between the typed and imputed genetic variants in our studied African infants and HLA allele diversity across the African continent. HLA alleles are known to vary across populations and there has traditionally been a bias towards cataloguing class I allele diversity owing to recognised associations with multiple traits including malaria and HIV. We therefore performed high resolution typing for three class I and eight class II *HLA* genes in a total of 1,706 individuals from African and admixed African-American populations. 832 individuals were included from the 3 *VaccGene* populations, alongside 634 individuals from 6 African populations (Esan in Nigeria (ESN), Gambian in Western Division, The Gambia – Mandinka (GWD), Luhya in Webuye, Kenya (LWK), Maasai in Kinyawa, Kenya (MKK), Mende in Sierra Leone (MSL), and Yoruba in Ibadan, Nigeria (YRI)) and 131 from 2 admixed African populations (African Caribbean in Barbados (ACB), and African Ancestry in Southwest USA (ASW)) from the 1000 Genomes project^21^. Newly sequenced individuals from the MKK population were included in this analysis with sample identifiers provided **Table S4**. With the exception of the new *VaccGene* populations and MKK individuals, all other individuals were selected on the basis of availability of DNA for classical HLA typing and whole-genome DNA variant calls available through genotype or whole-genome sequence data.

As summarised in **Fig 1C** (with a breakdown of numbers of individuals from each population with genotype, whole genome sequence, and diverse HLA type information available on each platform provided in **Table S5**), we employed three separate typing platforms to ensure the highest quality HLA allele calls, to protein coding level of resolution, possible for the continent. Our first objective was to ensure that any HLA calls derived from a short-read (MiSeq) next-generation sequencing platform was equivalent to traditional Sanger based typing, that has traditionally been considered the Gold Standard in clinical facilities. Using 47 randomly selected individuals from Uganda (discussed in **Supplementary Text**) we found all calls derived from Sanger based typing were also made using MiSeq and thus quality was considered equivalent. However, the ability to distinguish *cis*/*trans* strand state with the MiSeq platform reduced the number of potential ambiguous calls when two heterozygous alleles occurred in an individual and thus when considering the potential scalability and cost-effectiveness for large-scale typing we elected to proceed with MiSeq for the next stage of validation. Our second objective of this phase of the project was to determine the number of novel protein coding HLA alleles detectable in our tested African populations, some of which are historically poorly characterised. We used long-read PacBio technology to sequence exons of HLA genes in up to 836 individuals where MiSeq data was also available across all populations. With the exception of individuals from BF, all tested populations were found to possess at least one novel allele at one locus using one or other of the sequencing methods, although overall frequencies of novel allele detection were low, with less than 5% of all typed individuals possessing novel protein coding alleles detectable at any locus (**Fig 1D**). However, some populations did exhibit higher proportions of novel alleles than others with over 4% of MKK individuals possessing novel alleles detectable by either MiSeq or PacBio typing methods at HLA-A, HLA-DPA1 and HLA-DQB1 loci, and novel HLA-DPA1 alleles were detected in all except the West African BF and MSL populations. Overall, there was little advantage in applying PacBio to detect novel alleles compared to MiSeq for the purposes of novel protein coding allele detection and therefore MiSeq was used for all further downstream analyses. Together these results serve to highlight the importance of understanding the distribution of novel alleles in populations traditionally under-represented in genomic studies to date, especially in relation to complex regions of the genome such as HLA.

In order to understand allelic diversity in this dataset, and thus the importance of including representatives from all tested populations across the continent, we calculated pairwise population differentiation estimates (using G_ST_) between the tested populations using 6-digit ‘G’ coding of allelic variation (G_ST_ explicitly accounts for multi-allelic sites and is therefore preferred over F_ST_ in such scenarios). We noted some loci to be substantially differentiated across the continent, as already known, including HLA-B, HLA-C and HLA-DRB1 (**Fig. 1E**). However, we also noted that there was significant differentiation at the HLA-DPB1 locus with some estimates >0.5, equivalent to HLA-B, which has rarely been described in Africa and is even clearly observed at the lower 2-digit (1 field) level of resolution as shown in the pie-charts matched to population geography in **Fig. 1F**. However, most of the high levels of differentiation observed in HLA-DPB1 were linked with the MKK individuals who also appeared to have a preferential differentiation of HLA-C, and HLA-DP loci compared to other populations (**Fig. 1E**). Otherwise, differentiation was high (>0.4) for HLA-B, HLA-C and HLA-DRB1 loci in a non-specific population way supporting the inclusion of as many different continental populations as possible in the African HLA imputation reference panel.

### An HLA imputation reference panel for Africa

We next combined these high resolution 3-field (6-digit ‘G’) resolution HLA types derived from MiSeq with genotype data from 1,597 individuals across the same 11 African populations to generate a large, comprehensive HLA imputation reference panel available for African populations (**Fig. 2A**; see Data Availability in Methods). Variant calls across the region were available either from direct array genotyping or next-generation sequence (NGS) data. It is unclear whether differences in platform typing technology adversely affect imputation performance, therefore we first merged the variant calls determined using either dataset by only including variants that had a very high (r^2^>0.999) level of concordance between overlapping array and NGS calls. For this first validation step we elected to use HLA*IMP:02 for imputation given the explicit design to handle missing data and the reported high performance in populations of African descent^22^. We found that there was very little difference in allele concordance estimates between calls derived from either NGS or genotype in populations where we had both calls available (ACB, ASW and YRI) (**Fig. 2B**). Therefore we proceeded to build the imputation panel and algorithm based on HLA*IMP:02, using the merged genotype/NGS variant calls and accounting for higher resolution HLA allele calls. We called this new system HLA*IMP:02G. We then compared the performance of three algorithms for imputation compared to MiSeq typing as Gold Standard and using a five-fold cross-validation approach. The compared algorithms and reference panels were HLA*IMP:02G (the new system using MiSeq HLA calls and variant calls derived from genotyping and NGS), the original HLA*IMP:02 algorithm using a multi-ethnic reference panel, and a recently developed multi-ethnic imputation reference panel (the Broad multi-ethnic (ME) HLA panel)^23^. Only calls to 2-field (4-digit) resolution were available for HLA*IMP:02 and overall we observed a significant improvement in calling at all loci with the new HLA*IMP:02G algorithm compared to HLA*IMP:02 (**Fig. 2C** with performance statistics available in **Additional Tables 2 and 3**). The exceptions to this were HLA-A in Burkinabe individuals, as well as HLA-DRB4 and - DRB5 across all populations which are known to be minimally polymorphic. In keeping with our observation of increased differentiation at HLA-DP loci, we observed the greatest increase in performance for HLA-DPB1 where the mean concordance using HLA*IMP:02 was 0.42, increasing to 0.92 with HLA*IMP:02G. In contrast, for our comparison with the Broad ME-HLA panel we compared 6-digit ‘G’ resolution calls and although we still observed consistent improvements with HLA*IMP:02G, some alleles were called as effectively using the ME-HLA panel (such as HLA-A, HLA-B, and HLA-DRB1, **Fig. 2D** with statistics available in **Additional Table 4**). The most significant improvements between algorithms were again seen for HLA-DPB1 (mean with ME-HLA 0.74 *vs* 0.92), HLA-DPA1 (0.79 *vs* 0.97) and HLA-DQB1 (0.80 vs 0.96). These results support not only the inclusion of diverse populations in African-specific reference panels to substantially improve the performance of population-specific HLA allele imputation, but also highlight the benefit of targeted typing in some individuals to further refine population-specific signals. Our results also demonstrate that it is possible to incorporate genotype variants of differing technology backgrounds that may be used for imputation without adversely affecting imputation quality.

**Fig. 2.**
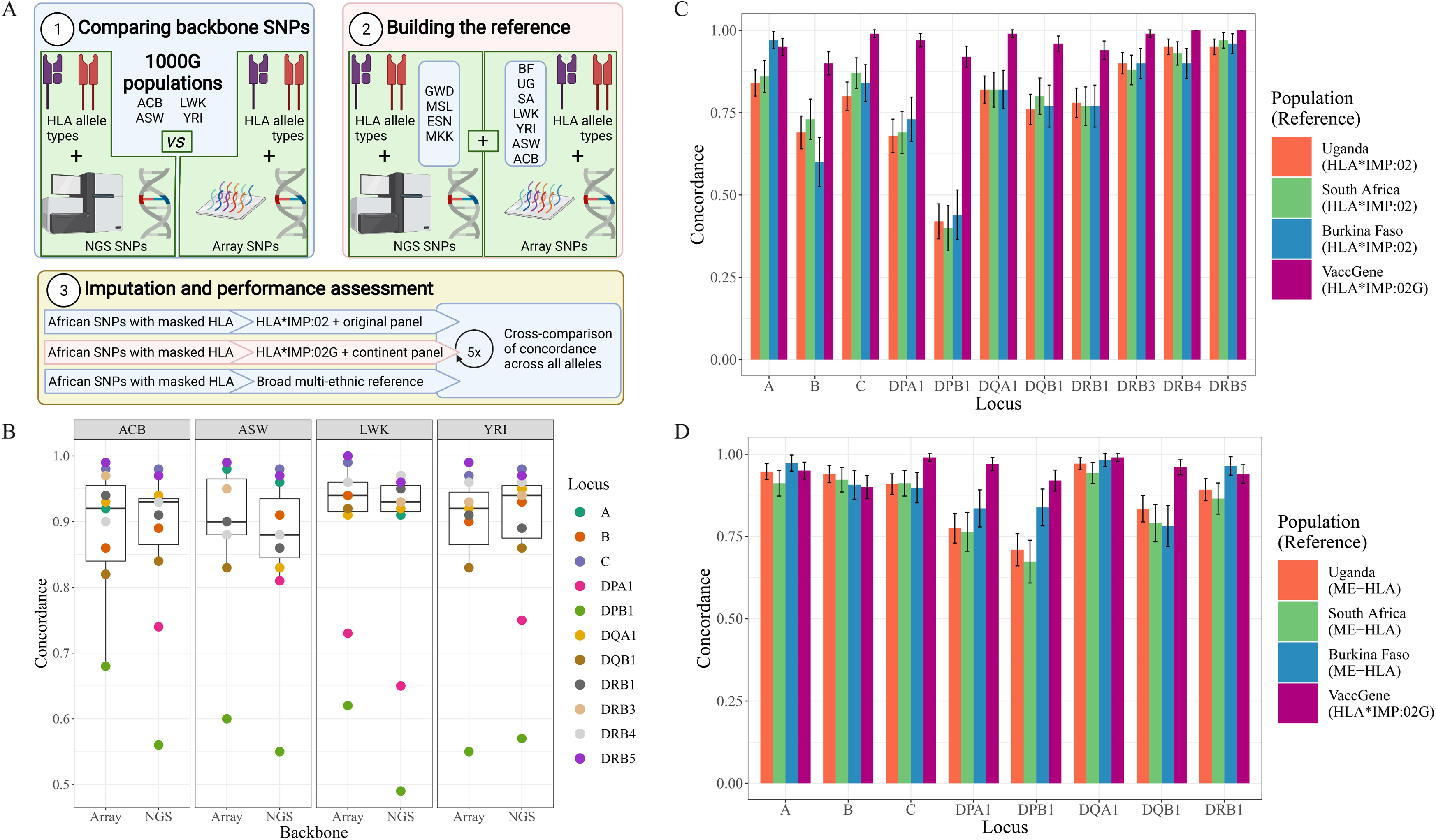
Imputing HLA alleles in African populations using a continental reference panel. **(A)** Schematic of approach to build and test a novel reference panel and adapted algorithm for imputation of HLA alleles in Africa. **(B)** The first stage involved testing for differences in imputation performance (using the original HLA*IMP:02 algorithm) with individuals from four African populations with variant data called by array genotyping or next-generation sequence data (NGS). Points are concordance estimates between imputed and MiSeq called HLA alleles for each gene locus. The box plot centre line represents the median; the box limits, the upper and lower quartiles; and the whiskers are the 1.5x interquartile range. **(C)** HLA imputation performance (measured as locus-specific concordance between alleles called to 2-field (4-digit) resolution) in the VaccGene populations using the traditional method and reference set (HLA*IMP:02) clustering by locus and population. Results are compared to the performance of our enhanced high-resolution algorithm and reference data-set (HLA*IMP:02G) using the same individuals divided into validation and test groups using a five-fold cross-validation approach. Means of performance and 95% confidence intervals are plotted for each comparison. Full statistics are available in **Additional Data Tables 2**, **3** and **4**. **(D)** HLA imputation performance comparing results from the Broad multi-ethnic reference panel to that from HLA*IMP:02G called to 6-digit ‘G’ resolution. ACB: African Caribbean in Barbados; ASW: African Ancestry in Southwest USA; LWK: Luhya in Webuye, Kenya; YRI: Yoruba in Ibadan, Nigeria.

### Fine-mapping HLA association results with vaccine antigen responses

We used our imputed HLA results to test for association between the 71,297 variants, 164 HLA alleles and 2,809 HLA amino acid residues with a minor allele frequency >0.01 before employing step-wise fine-mapping to identify 12 statistically significant (*P*_pooled_ ≤ 5 x 10^-9^) novel associations with each of the vaccine traits mapping to multiple HLA class II loci. Stepwise conditional regression results are shown in **Figs. S2A-S2C** and the final results after a combination of manual and automated regression modelling are provided in **Fig. 3** with the statistics provided in **Table S7** and with evidence of heterogeneity provided in **Table S8**. We observed that each of the traits exhibited multiple, independent association signals that were best explained by either HLA alleles, SNPs or amino acids each in different HLA genes. For diphtheria, for example, we found that the same SNP as identified in the first round of analysis (rs34951355) provided the smallest *P*-value and explained the association most parsimoniously. In contrast, PT was best explained by two independent associations: the same SNP as identified in the genotype-only GWAS (rs73727916), and the presence of the amino acid glutamine at position 74 of HLA-DRB3 (DRB3-Gln, beta_univariate_=-0.31, *P*_univariate_=4.2×10^-25^) which exhibited effects in opposite directions. The FHA association was best explained by two HLA alleles (HLA-DRB1*15:03:01G and HLA-DRB1*08:04:01), whereas both PRN and HBsAg were explained by four independent associations spanning HLA-DRB1, and HLA-DQ and HLA-DP amino acids respectively. For those primary associations where there was little evidence of heterogeneity we found that individuals carrying HLA-DRB1*08:04:01 had 1.5x greater FHA antibody levels than those who did not carry this allele (geometric mean titre 6.30 EU/ml (95% confidence interval 5.14-7.73) compared to 4.24 EU/ml (4.04-4.46)). We also observed that individuals carrying HLA-DRB1*11:02:01 had 1.8x greater PRN antibody levels than those who did not (22.98 EU/ml (17.31-30.51) vs 12.97 EU/ml (12.27-13.71)), and individuals carrying DRB1-74Arg had 0.6x less HBsAg antibody than those not carrying the allele (69.21 mIU/ml (50.94-94.21) vs 106.84 mIU/ml (97.48-117.09)).

**Fig. 3.**
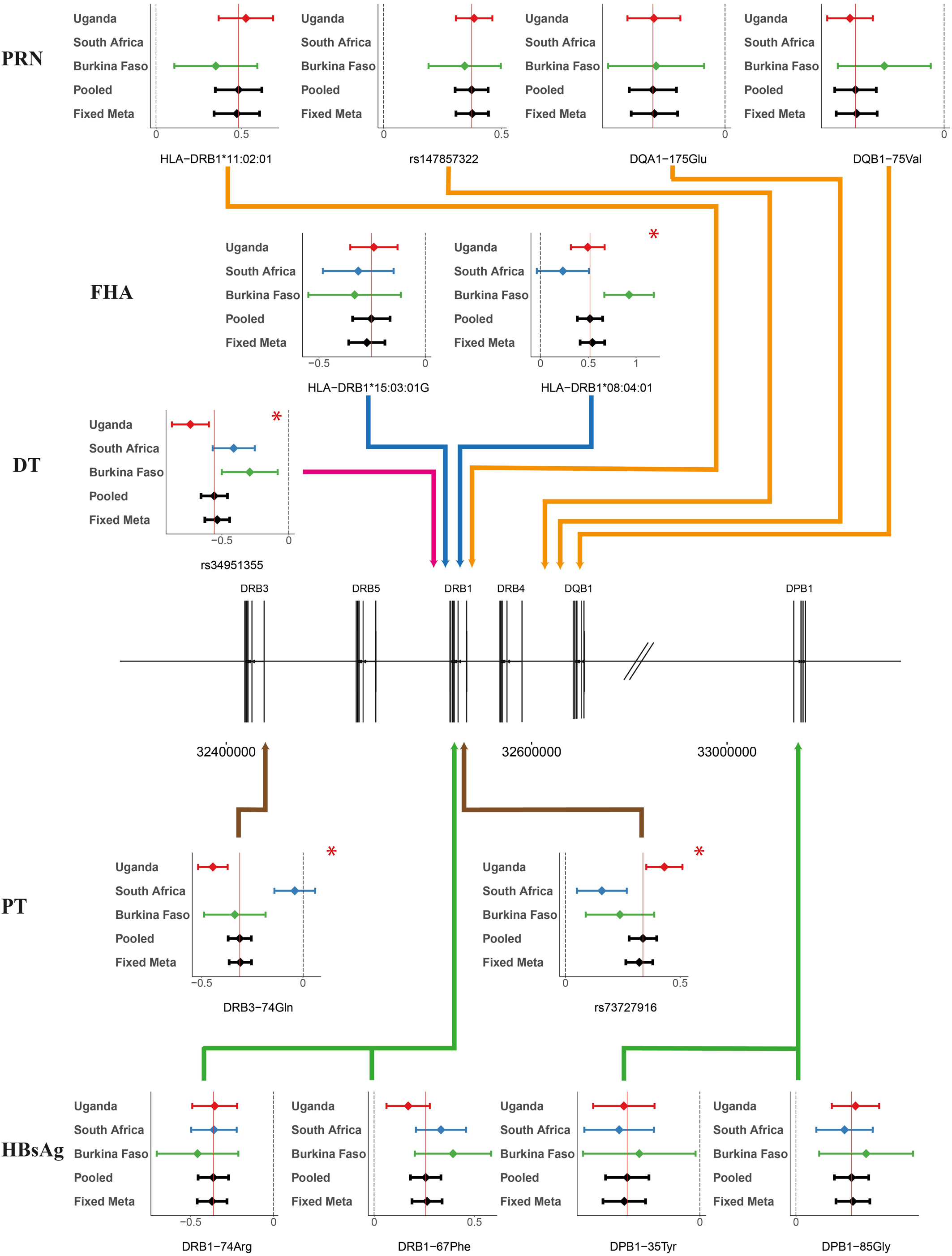
HLA associations with vaccine responses fine-mapped to HLA variants. Forest plots of effect estimates (points) for fine-mapped variants for each trait colored by population (Uganda as red, South Africa blue and Burkina Faso green) with 95% confidence intervals (bars) and corresponding distributions for the pooled linear mixed model (‘Pooled’ – solid black horizontal line) and fixed effects meta-analyses (‘Fixed Meta’). Variants were deemed to be independently associated with each trait using combined manual and automated regression approaches. Dashed vertical black lines represent no effect (beta=0) and solid vertical red lines cross the beta estimate of the Pooled model as a reference. The originating locus of association is represented by solid arrowed lines colored by trait indicating the relevant region of association on chromosome 6. Associations demonstrating significant evidence (*P*Q ≤ 1×10^-3^) of heterogeneity are highlighted with a red asterisk (*). Pertactin was not administered to South African infants hence there are no measured effects for this population. PT: pertussis toxin, FHA: pertussis filamentous hemagglutinin; PRN: pertussis pertactin; DT: diphtheria toxin; HBsAg, hepatitis B surface antigen.

To put our association findings in the context of public health we used other data available from the African infants to understand the impact of genetic variation on vaccine immunogenicity compared to other important variables available from our datasets. We explored the proportion of variance explained by variables including time between vaccination and sampling (included as a covariate in all GWAS models), sex, weight-for-length z-score at birth, and HIV status for each cohort and vaccine response where available, and compared these to the proportion of variance explained by the HLA genetic variants for each antibody trait (**Fig. 4A**). We found that the contribution of genetic associations consistently outweighed the impact of other variables except that of the time between vaccination and sampling. Overall we observed little effect of sex or weight-for-length on the variance when measured at the time in our study, and although the proportion of variance explained by HIV status across each of the populations was minimal, the small number of individuals infected with HIV at birth in Uganda did have significantly lower levels of antibody against all tested vaccine responses with the exception of FHA (**Fig. 4B**). The mean proportion of variance explained by the HLA variants across the three tested populations was 5.7% (range 1.5%-10.9%) for PT, 6.1% (1.6%-13.8%) for FHA, 10.4% (9.3%-11.4%) for PRN, 4.3% (1.2%-7.0%) for DT and % (5.2%-9.1%) for HBsAg emphasising the importance of genetics impacting overall response to multiple vaccines in infancy.

**Fig. 4.**
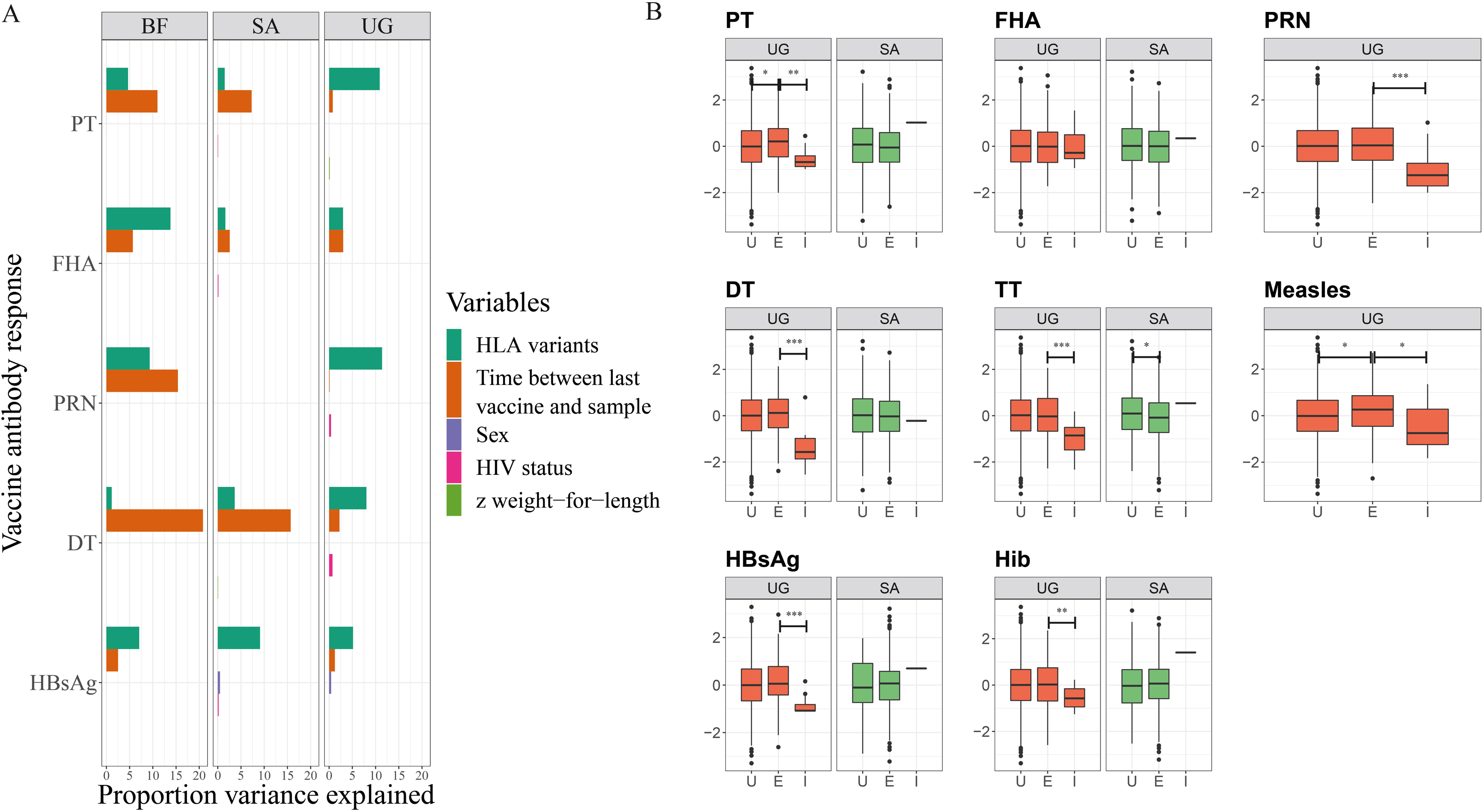
Assessing the impact of genetics and other exposures on magnitude of vaccine response in VaccGene. **(A)** The proportion of variance explained (r^2^) by genetic variants (those fine mapped to be most relevant as in Fig 3 for each antibody trait), time in weeks between last vaccine and sampling for antibody assay, sex (male vs female), HIV status (uninfected (U), exposed (E) or infected (I) at birth) and z weight-for-length score at birth, were available in each tested cohort. **(B)** Distributions of antibody responses stratified by HIV status at birth in Ugandan (UG) and South African (SA) individuals with differences tested between strata using the Wilcoxon rank test. The box plot centre line represents the median; the box limits, the upper and lower quartiles; and the whiskers are the 1.5x interquartile range. * *P*<0.05; ** *P*<0.01; *** *P*<0.001.

### Correlating vaccine immunogenicity and effectiveness through genetic associations

Given the observed impact of genetic variants on antibody response, we next aimed to understand these genetic associations in the context of vaccine effectiveness. Genetic analyses of cohorts of vaccine failures are rarely available, largely attributable to the success of vaccines and the challenges in identifying, recruiting and sampling individuals with recorded vaccine failure. A large independent case-control genetic association study of self-reported pertussis (defined as the characteristic whooping cough) is, however, available and was undertaken using data from vaccinated adolescents and young adults in the United Kingdom who had received pertussis vaccine^24^. Comparing our pertussis antigen vaccination genetic association results to those from this pertussis GWAS, we found strong evidence of a negative correlation between the effect estimates for both SNPs (**Fig. S3A**) and amino acid residues (**Fig. 5A**) on antibody responses to PT, and susceptibility to pertussis (for amino acid residues, where more complete data were available, Pearson’s r=-0.83, *P_perm_*<1×10^-8^ after 10^8^ permutations (**Fig. 5B**)). No such correlation was observed for either SNPs (**Figs. S3B** and **S3C**) or amino acid residues (**Figs. S3D-S3G**) in association testing with the other two pertussis antigen responses in our study: PRN (amino acid r=-0.02, *P*_perm_ =0.57) or FHA (amino acid r=-0.01; *P*_perm_ =0.91). The observed amino acid correlation persisted after stringent correction for LD (**Fig. S3H**).

**Fig. 5.**
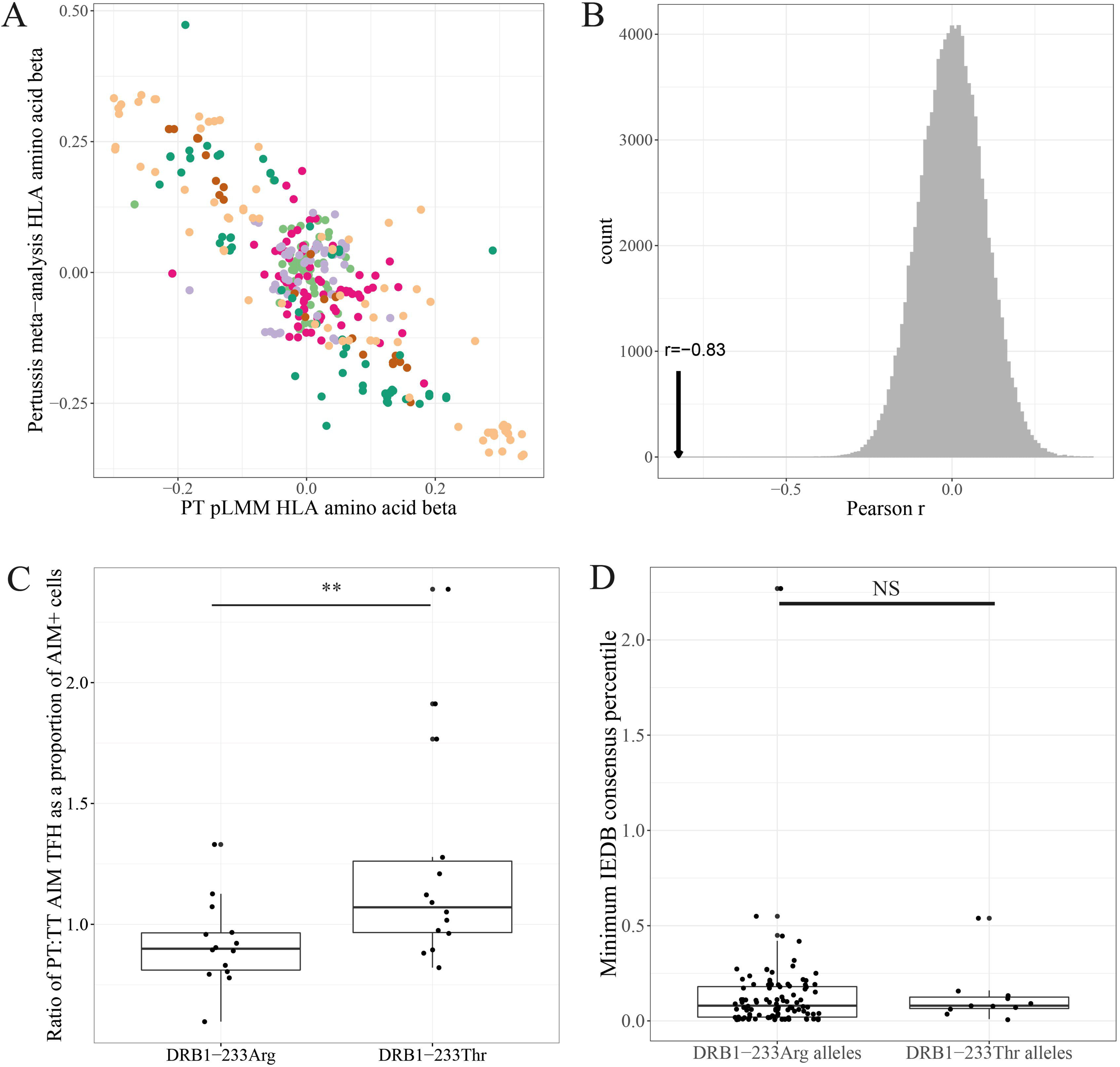
Mechanisms associated with HLA-mediated responses and vaccine failure. (A) The beta effect estimates for association between HLA amino acid residues and PT antibody response in the VaccGene infants are plotted against the equivalent estimates from a case-control association study of self-reported pertussis. Residues are colored by HLA gene (light green HLA-A; rose HLA-B; lavender HLA-C, orange HLA-DQA1; dark green HLA-DQB1 and gold HLA-DRB1). (B) Distributions of Pearson’s r coefficient following 100,000 permutations to measure the significance of correlation between effect estimates of HLA amino acids pruned by LD comparing responses against PT and against the pertussis GWAS. Pearson correlation coefficients were calculated after relabelling of the whooping cough GWAS variants generating the null distribution. The correlation coefficients determined using the true datasets are represented with a vertical arrow. (C) Ratio of circulating pertussis:tetanus toxin (PT:TT) specific T_FH_ in donors of known HLA-DRB1 type divided by the index HLA-DRB1 variant associated with PT antibody response and pertussis self-report. Antigen-specific T_FH_ cells are represented as a proportion of all cells categorized as Antigen Inducible Marker (AIM+) cells. (D) Predicted affinities for top PT-derived peptides predicted to bind to alleles with those containing a threonine at position 233 of HLA-DRB1 (‘DRB1-233Thr’) compared to those with an arginine (‘DRB1-233Arg’) calculated from the immune epitope database. The box plot center line represents the median; the box limits, the upper and lower quartiles; and the whiskers are the 1.5x interquartile range. ** *P*<0.01; NS not significant

Since the majority of participants in the UK-based pertussis analysis were likely to have received a pertussis vaccine, these data provide evidence that i) both PT-specific antibody responses and risk of post-vaccination pertussis exhibit significant associations with genetic variation, ii) the genetic architecture of PT responses and pertussis are negatively correlated and thus iii) it is likely that PT is a key correlate of efficacy in pertussis and iv) these effects are consistent across populations of diverse ancestry. Although the variants identified as most relevant for PT in our study in African children were not all available in the pertussis study, the most significantly associated risk variant in the pertussis analysis (an arginine at position 233 in HLA-DRB1) had an odds ratio of 1.38. The same variant alone accounts for 6.1% of variance of PT antibody response in the UG cohort demonstrating the potential importance of genetic variation on both antigen immunogenicity and vaccine effectiveness. This allele is common, with a frequency of 35% of the UK population, and 48% in our tested African populations suggesting that, if confirmed, the effects could be significant in most populations of the world.

### Testing effects of HLA associations on follicular-helper T-cells

In comparison to autoimmune conditions where HLA associations are recognised but the driving antigens are less well defined, our observed HLA associations with vaccine responses offer the opportunity to explore the underlying mechanisms of genetic associations given the explicit knowledge of driving antigens. We first sought to test whether we could confirm the observed association between HLA and PT response in an independent cohort and whether we could provide evidence that this effect persisted through the relevant antigen presentation-T cell axis. To achieve this, we elected to use a genetic variant that was known to affect both PT response and pertussis susceptibility and would be readily available through HLA typing. However, we had to decide between an HLA-DRB3 variant that was most associated in our antibody analysis but was not present in the published analysis of pertussis, and an HLA-DRB1 variant that was both typed and found significantly associated with the tested traits in both studies. We therefore accessed a component of the individual-level pertussis GWAS data (Avon Longitudinal Study of Parents and Children; ALSPAC) and performed dedicated imputation of HLA-DRB3 in this cohort. We found that although a negative correlation was still observed across HLA-DRB1 amino acids in this cohort (r=-0.55, P_perm_<1×10^-5^), there was no such signal across HLA-DRB3 (r=0.13, P_perm_=0.16). Thus, allowing for the assumption that the genetic architectures of PT response and pertussis susceptibility are linked functionally, these results from our multi-ethnic multi-phenotype analyses suggest that the functional variant is most likely to reside in HLA-DRB1. The most significantly associated HLA-DRB1 variant in both studies is the aforementioned position 233, which may be either an arginine (DRB1-233Arg) as described earlier, or a threonine (DRB1-233Thr). Arginine is found in this position in alleles such as HLA-DRB1*11:02:01 (P_pooled_=3.2×10^-7^, beta −0.32, SE 0.06 from our African vaccine GWAS of PT response) and the threonine in allele groups such as HLA-DRB1*15:03:01G, (*P*_pooled_=4.3×10^-11^, beta 0.30, SE 0.05), associated with lower and higher antibody responses respectively. We therefore stratified individuals from an independently recruited set of individual from studies in the United States (hereafter referred to as the ‘Sette studies’) into two groups based on whether they carried an arginine or a threonine at this position 233 in HLA-DRB1. We compared levels of antigen-specific follicular-helper T-cells (T_FH_)^25^ between individuals in the Sette studies homozygous for alleles encoding either residue at this HLA-DRB1 position (**Fig. S3I and Table S9**). We found that individuals carrying a threonine had, on average, a 1.2 fold greater ratio of pertussis:tetanus toxin specific T_FH_ compared to individuals carrying arginine (one-tailed Mann-Whitney *P*=0.007; **Fig. 5C**). Despite these associations, we found no evidence of differences in the affinity (**Fig. 5D**) or breadth (**Table S10**) of PT peptide binding defined by residues at position 233 of HLA-DRB1 using *in silico* peptide-binding methods. Thus, these data provide evidence in favor of the T_FH_-B cell axis being a key pathway involved in differential pertussis vaccine response and protective efficacy mediated through the HLA-DRB1 locus although these data go against the model of improved antigen-specific peptide binding driving these effects.

### HLA expression quantitative trait loci in Africa correlating with vaccine responses

Given, firstly, the observations that, for PT, HLA binding may not be the predominant mechanism driving an activation of antigen-specific T-cells, and secondly, for DT, the signal was almost exclusively explained by a SNP (rs34951355) alone with no obvious link to peptide-binding, we next aimed to test the hypothesis that HLA gene expression may play a role in driving these traits. We developed two expression quantitative trait loci (eQTL) resources to test this hypothesis. The first resource was designed as a well-powered tool, representative of African population immune cells. We combined available HLA-wide genotypes with RNA sequence data derived from immortalized lymphoblastoid cell lines from many of the same individuals included from our imputation reference panel from 1000Gp3 (n=655 from 6 African populations with the significance of SNPs on *cis*-expression of genes provided in **Fig. 6A** and **Additional Data Table 5**). Such an analysis has traditionally been challenging owing to difficulty mapping polymorphic reads to a single European ancestry reference genome but our method of using a personalized reference sequence with high resolution data allowed a sensitive detection of eQTLs across 4 genes in particular: *HLA-A, HLA-C*, *HLA-DRB1* and *HLA-DPB1*. Secondly, to allow an improved understanding of the cell-specific impact of variants we applied the same bioinformatics pipeline to a published *ex vivo* cell-specific eQTL dataset^26^ including 13 cell types (naïve and activated lymphocytes and monocytes and NK cells). Inspecting the correlation between *P-* values for variants modulating expression of *HLA-DRB1* between cell types (those with – log_10_(*P*)≥3, **Fig 6B**) we see a high level of correlation for some cell types (stimulated CD4 and CD8 T-cells rho 0.93, and monocytes and naïve B-cells rho 0.78 as examples), whereas for others the correlation was poor (monocytes and NK cells rho −0.12). Using these datasets, we first inspected the DT associated variant which was a nucleotide substitution located within intron 1 of *HLA-DRB1* with the minor allele associated with reduced DT antibody levels. The index variant itself was not called with high confidence across all populations in our eQTL datasets, and therefore we assessed the impact of another variant in LD (rs545690952, r^2^=0.80 located in intron 2 of *HLA-DRB1, P_pooled_*= 3.0×10^-27^, beta=-0.49, SE=0.05 from the African infant DT GWAS) on expression of *HLA* transcripts. We found that the alternate guanine allele of rs545690952 was associated with statistically significant downregulated expression of *HLA-DRB1* (*P*_meta_=1.6×10^-4^, **Fig. 6C**) and *HLA-DQB1* (*P*_meta_=3.9 x 10^-5^) suggesting that variation in DT response may be mediated by changes in *HLA* gene expression. In the cell specific datasets, we found the only significant effect of rs545690952 on *HLA-DRB1* expression was in monocytes in the same direction (*P*=6.3×10^-3^, **Fig. 6D**) consistent with a cell-specific effect in one of the most critical antigen presenting cells present in the circulation. A non-significant trend of association in the same direction was observed with naïve B-cells which is consistent with our observed signature correlations, the derivation of lymphoblastoid cells lines from B-cells, and the known antigen presentation ability of this cellular subset.

**Fig. 6.**
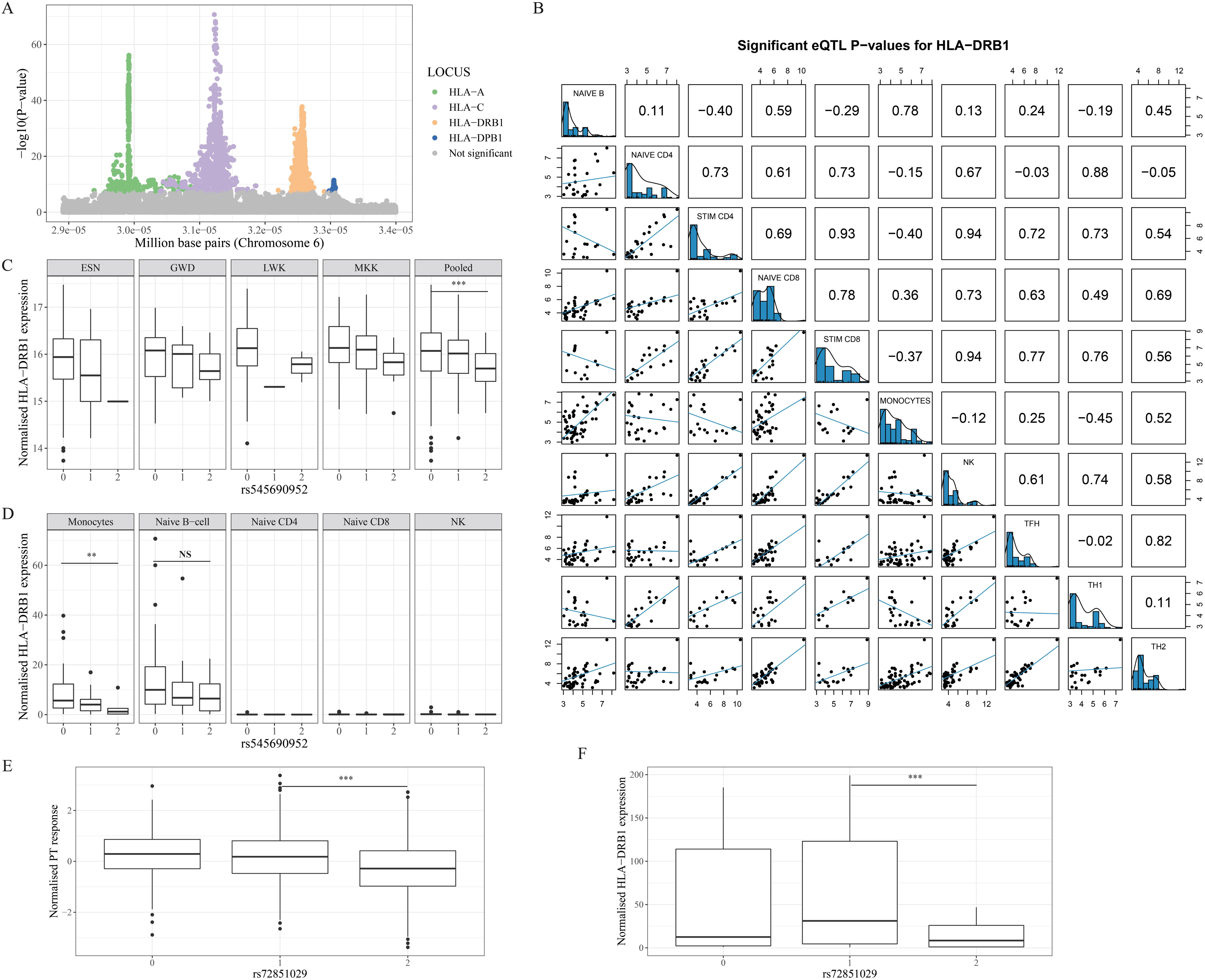
Mapping cis-eQTLs across the HLA in diverse immune cells. **(A)** Variants with evidence of being *cis*-expression quantitative trait modulators are plotted by position across the HLA against evidence of significance of impacting expression of four *HLA* transcripts. Only variants with significant evidence (*P <* 5 x 10^-8^) are colored by gene with the remainder in grey. RNA sequence data from lymphoblastoid cell lines were mapped to personalized *HLA* gene sequences derived from high-resolution typing. **(B)** The correlation in *P*-value estimates for variants predicted to be cis-eQTL variants in different cell types from the DICE dataset. 10 of 13 cell types are presented with scatter plots in the lower half of the table and Spearman rho estimates in the upper half. **(C)** Effect of a variant in LD with the index DT-associated variant on levels of *HLA-DRB1* in four populations (ESN, GWD, LWK, MKK) with more than a single observation in each genotype category. A plot of the data from the pooled set of four populations is shown for each gene. The x-axes numbers refer to the number of copies of the minor G allele compared to the major T in each group of individuals per population. **(D)** The effect of this same variant on *HLA-DRB1* expression in circulating monocytes, naïve B-cells, naïve CD4 and CD8 T-cells and natural killer (NK) cells from the DICE study demonstrating a consistent direction of effect in monocytes. **(E)** The effect of alternate T alleles of rs72851029 on PT antibody response in the African infant GWAS with significance tested in a recessive model. **(F)** The effect of rs72851029 on *HLA-DRB1* expression in monocytes with significance tested using a recessive model. The box plot center line represents the median; the box limits, the upper and lower quartiles; and the whiskers are the 1.5x interquartile range. ** *P* < 0.01, *** *P*<0.001, NS: not significant.

For PT, we aimed to test the hypothesis of gene expression in the independent peak that we had shown earlier was associated with T-cell activation in the absence of binding effects and where HLA-DRB3 was unlikely to play a functional role. In the cluster of associated variants, the nucleotide most associated with PT was rs72851029 (*P*_pooled_=6.6×10^-25^) where the alternate thymine allele was associated with decreased PT antibody response (**Fig. 6E**), decreased *HLA-DRB1* expression in the African lymphoblastoid cell lines (*P*_meta_=1.25×10^-22^) and decreased *HLA-DRB1* expression in monocytes in our cell-specific analysis in pattern consistent with a recessive inheritance (*P*=5.0×10^-4^ **Fig. 6F**). Altogether these data provide further evidence that *HLA-DRB1* expression may play a major role in influencing pertussis and diphtheria antibody responses, as well as potentially in risk of pertussis following vaccination with acellular pertussis vaccine.

## Discussion

Vaccines are one of the most successful public health interventions of the modern era. Despite their effectiveness spanning multiple infectious diseases, many challenges remain in ensuring their continued success. Exemplar challenges include understanding the mechanisms of breakthrough infections occurring despite vaccination, following pertussis vaccination for example, in addition to the challenges with developing vaccines against infections including TB and HIV. Here we investigated the impact of human genetic variation on vaccine immunogenicity and effectiveness for key vaccines integral to the EPI in African infants. We found that genetic variation across the HLA is strongly associated with variable antibody responses against five of the eight vaccine antigens measured in our study. We then developed a dedicated HLA imputation resource using accurate high-resolution MiSeq typing and fine mapped the signals of association to a variety of HLA variants and alleles. Using a variety of approaches we found evidence that variants in HLA-DRB1 are associated with increased PT-specific T_FH_ activity and, thus, in turn increased antibody production and ultimately protection against whooping cough. However, we found less evidence of an effect mediated through predicted binding but instead, more evidence of an effect mediated through *HLA* gene expression, which was also found for DT antibody responses.

Together, our results provide substantial evidence of an influence of human genetic variation on multiple vaccines delivered to infants worldwide that until now have only been appreciated reproducibly for vaccinations targeting hepatitis B ^27, 28^, meningitis C^11^ and measles^29^, although only hepatitis B has well characterised associations across the HLA. The mechanisms underlying such associations have always been elusive and traditionally have been suspected to be predominantly driven by peptide binding^30^. To attempt to understand potential mechanisms in more detail we typed HLA alleles in as many individuals as possible to improve confidence in direct allele calling and downstream imputation in African populations. Although we observed a significant level of novelty in protein coding alleles we did not observe these to occur at levels greater than 5% meaning that imputation and association testing for common alleles was still an appropriate method of analysis. Overall, however, given the significant differentiation of alleles across the continent, there remained substantial benefit to including individuals from as many populations as possible to improve imputation performance. As was expected by using allele calls derived within our test dataset, the performance of imputation using our HLA*IMP:02G algorithm and reference panel was excellent, however it is worthwhile to note that a newly available imputation resource^23^ performed equivalently at multiple loci of anticipated medical importance.

Having access to the high-resolution HLA calls not only had benefits for imputation and fine-mapping the associated variants, but also for generating high confidence calls of eQTLs across the locus for *HLA* genes. Differential expression of *HLA-C* has been linked with susceptibility to HIV disease progression but there are limited datasets available for characterising HLA expression at multiple points across the locus. Our multi-population, personalised, multi-gene and multi-cell type HLA eQTL resource highlights the potential importance of this mechanism for vaccine responses that may act on its own or synergistically alongside peptide binding or other peptide processing defects in a number of traits as is already being recognised in autoimmunity^31^.

The clinical relevance of our work is multi-fold. Firstly, if further shown to be true, our results would suggest that expression of *HLA* genes may be a significant driver in differential vaccine response. Adjuvantation is well recognised to boost immune responses that may in part be due to increased expression of *HLA* genes^32^ but the cell-specific effect of such methods are poorly characterised. It may be that more appropriate targeting of adjuvantation for vaccines such as pertussis may help boost universal protection and reduce risks of breakthrough. Secondly, although population scale differences are unlikely with pertussis (because the frequencies of the linked alleles in UK and African populations were very similar), it is highly plausible that HLA associations could have greater relevance for some populations more than others. Risks of breakthrough infection may be more common in some populations owing to genetic differences and thus consideration of these differences may be important for future vaccine delivery. Finally, if the impact of genetic variation on the effectiveness of vaccination was higher for vaccines other than pertussis or diphtheria (HIV for example), then it would be even more important to identify these associations *a priori* before making statements about individual level, or population-scale vaccine effectiveness.

The potential limitations of our work include the varied nature of both the methods used for HLA typing or inference and the heterogeneous nature of the cohorts used for the vaccine response genetic association studies, which could all affect the interpretation of our results. We explicitly designed the study to allow cross-correlation between HLA allele calls defined by Sanger sequence, short-read MiSeq and long-read PacBio sequencing methods. Even in these relatively understudied populations our results are in agreement with all work undertaken in other populations demonstrating that most inconsistencies between platforms would be explained by differential exon coverage and that when described to exonic sequence level, most alleles had already been reported. Thus, the short-read MiSeq offered the most cost-effective scalable method to type large numbers of individuals to a consistent standard. Given the possibility of expression effects modulating functional responses, the future exploration of intronic variants, which are more likely to directly regulate expression, will be substantially improved by long-read sequencing technologies. As reference databases accumulate more long-range sequences, the full contribution of coding and non-coding variants to downstream functional effects will become more apparent. Our findings highlight the importance of the HLA-DP locus in particular. These were not only observed to be significantly differentiated worldwide, but were also found to be significantly associated with HBsAg in line with several previous reports^27, 33, 34^. Together with increasing reports of an HLA-DP association with other viral infections including SARS-CoV2^35^, these results highlight the growing importance of understanding the diversity and cellular function of this locus in multiple populations. Finally, although the cohorts included in the vaccine response GWAS were selected to represent diverse geographical and environmental exposure backgrounds, many of the effect estimate signals were remarkably homogeneous with the best example being the HLA-DRB1 signals observed for HBsAg. Significant heterogeneity was observed for some association signals including the index HLA-DR signal observed with PT where a null association was observed for the SA cohort. This absence of association could be related to the use of an acellular as opposed to a whole-cell vaccine in South Africa which is the only obvious difference in vaccine delivery, or could be as a result of a yet unidentified genetic or other population cause of heterogeneity. These issues also highlight the ongoing challenges with reliably fine-mapping association signals clearly across such diverse populations. As demonstrated for pertussis, the most likely causal variant from our *VaccGene* cohort statistically was an HLA-DRB3 amino acid residue, but, when combining our data with that of a related phenotype from a UK dataset, we found near-equivalent evidence that the signal was instead linked to an HLA-DRB1 variant that could equally alter peptide binding or gene expression. Given many acknowledged challenges of fine mapping in this complex locus, our work demonstrates that further understanding will only come from improved resource availability and a multiplicity of technical approaches to reliably pin-point the underlying mechanism.

In conclusion, our results demonstrate that variation of HLA gene expression is likely to play a role as part of a multi-faceted set of mechanisms influencing important biological processes. Resources such as our collective African genetic and transcriptomic datasets may be key to understanding multiple genetic associations across the HLA with traits of importance across Africa within a functional context.

## Data Availability

Summary statistics are available as part of the submission or on Zonodo (https://doi.org/10.5281/zenodo.7357687).
Direct genotypes will be available through managed access through the European Genome-Phenome Archive under accession EGAS00001000918.

## Acknowledgments

We thank all sample donors who contributed to this study and staff involved in consenting, sample and data collection and preparation includes interviewers, computer and laboratory technicians, clerical workers, research scientists, volunteers, managers, receptionists and nurses.

## Funding

This project has received funding from the European Research Council (ERC) under the European Union’s [Seventh Framework Programme (FP7/2007-2013)] (grant agreement No 294557).

This work was supported by the Wellcome Trust grant numbers 064693, 079110, 095778, 217065, 202802 and 098051.

AJM was supported by an Oxford University Clinical Academic School Transitional Fellowship and a Wellcome Trust Clinical Research Training Fellowship reference 106289/Z/14/Z and the National Institute for Health Research (NIHR) Oxford Biomedical Research Centre (BRC).

NJT is a Wellcome Trust Investigator (202802/Z/16/Z), is the PI of the Avon Longitudinal Study of Parents and Children (MRC & WT 217065/Z/19/Z), and is supported by the University of Bristol NIHR Biomedical Research Centre (BRC-1215-2001), the MRC Integrative Epidemiology Unit (MC_UU_00011) and works within the CRUK Integrative Cancer Epidemiology Programme (C18281/A19169).

Computation used the BMRC facility, a joint development between the Wellcome Centre for Human Genetics and the Big Data Institute supported by the NIHR Oxford BRC.

Financial support was provided by the Wellcome Trust Core Award Grant Number 203141/Z/16/Z.

Computational support and infrastructure was also provided by the “Centre for Information and Media Technology” (ZIM) at the University of Düsseldorf (Germany).

The UK Medical Research Council and Wellcome (Grant ref: 217065/Z/19/Z) and the University of Bristol provide core support for ALSPAC.

This publication is the work of the authors and NJT will serve as guarantor for the contents of this paper. GWAS data for ALSPAC was generated by Sample Logistics and Genotyping Facilities at Wellcome Sanger Institute and LabCorp (Laboratory Corporation of America) using support from 23andMe.The cellular immunology studies were supported by National Institute of Health grants U19 AI118626 (AS) and U01 AI141995 (AS/BP).

The views expressed are those of the author(s) and not necessarily those of the NHS, the NIHR or the Department of Health.

## Author contributions

Conceptualization: AJM, DG, BP, AS, RN, AME, GM, AVSH, MSS

Methodology: AJM, DG, BP, AS, RN, AME, GM, AVSH, MSS

Analyses: AJM, ATD, MP, DG, DB, EK, TC, RdSA, SP, GS, SW, HK, CSLA, AR, DK, TP, KA, KE, TM. KE, NE, SP

Resource generation and data curation: AJM, MP, DG, TC, AM, CC, AD, HK, CP, NC

Funding and Supervision: AJM, KJ, FRMvdK, PK, BP, AS, NC, RN, SS, SM, AME, GM, AVSH, MSS

Writing—original draft: AJM, ATD, MP, DG, DB, EK, TC, GM, AVSH, MSS

Writing—review & editing: all authors

## Competing interests

Authors declare no competing interests.

## Data and materials availability

All data are available in the main text or the Supplementary Materials or in the European Genome-Phenome Archive under accession: EGAS00001000918.

## Materials and Methods

### Experimental Design and Study populations

The objectives of this study were to 1) test for association between genetic variation and antibody response to eight vaccine antigens delivered in infancy, 2) characterise the major *HLA* genes in a large collection of African populations using a range of sequence technologies, 3) use this resource to develop and test a population-specific HLA imputation panel, 4) use the high-resolution characterization to understand the likely functional mechanisms underlying these measured vaccine responses. The African populations included in this study include seven populations characterized as part of the 1000 Genomes phase 3 (1000Gp3) project, the Maasai from the HapMap collection, and three other populations recruited as part of the *VaccGene* initiative. The analyses used genotype data, described in more detail below, derived from array-based and / or next-generation sequence data alongside HLA allele information for all included populations. Association analyses were undertaken using only *VaccGene* populations incorporating array-derived genotype data alongside HLA allele types, vaccine antibody responses and clinical demographic data.

#### 1000 Genomes Phase 3 and HapMap Collections

The collection, genotyping and sequencing of the seven 1000Gp3 African populations have already been described *(36*) and all data are publically available (http://www.internationalgenome.org/). These populations include individuals from African Caribbeans in Barbados (ACB), Americans of African Ancestry in Southwest USA (ASW), Esan in Nigeria (ESN), Gambian in Western Divisions in the Gambia (GWD) of Mandinka ethnicity, Luhya in Webuye, Kenya (LWK), Mende in Sierra Leone (MSL) and Yoruba in Ibadan, Nigeria (YRI)). DNA was extracted from samples of publically available immortalized lymphoblastoid cell lines (LCLs) selected from unrelated individuals from these 1000Gp3 populations and from the Maasai in Kinyawa, Kenya (MKK) derived from the HapMap project^37^. The resultant DNA was used for short and long read HLA gene sequencing and typing. DNA from the MKK was also sequenced across the genome using short-read sequencing with all methods described in further detail below.

#### VaccGene populations

Participants included in the *VaccGene* study were recruited from three African countries selected partly due to their geographic dispersal across the continent and partly due the availability of high quality metadata and biological samples relevant to infant vaccination. These sites were in Uganda, South Africa and Burkina Faso. Individuals from each of the cohorts were included if their dates of birth, vaccination and blood sampling were available and if it was confirmed that they had received three doses of vaccines including diphtheria toxin (DT), tetanus toxin (TT), pertussis antigens, *Haemophilus influenzae* (Hib), and hepatitis B surface antigen (HBsAg) and a single dose of measles virus (MV) vaccine. The receipt of vaccines was confirmed through referencing the vaccination cards of infant participants or documented administration of vaccines by the research teams where relevant. Beyond exclusion criteria involved in preliminary recruitment of the individuals, no further exclusion occurred based on gender, ethnicity, HIV exposure or any other health status. A range of clinical and demographic metadata were collected from the three cohorts including the number of illnesses during the first year of life, details regarding the pregnancy and parental occupations and self-reported ethnicities (**Table S1**). A more detailed description of each of these populations follows below.

##### Uganda: The Entebbe Mother and Baby Study (EMaBS)

EMaBS is a prospective birth cohort that was originally designed as a randomized controlled trial to test whether anthelminthic treatment during pregnancy and early infancy was associated with differential response to vaccination or incidence of infections such as pneumonia, diarrhea or malaria (http://emabs.lshtm.ac.uk/)^38^. EMaBS originally recruited 2,507 women between 2003 and 2006; 2,345 livebirths were documented and 2,115 children were still enrolled at 1 year of age. Pregnant women in the second or third trimester were enrolled at Entebbe Hospital antenatal clinic if they were resident in the study area, planning to deliver in the hospital, willing to know their HIV status and willing to take part in the study. They were excluded if they had evidence of possible helminth-induced pathology (severe anemia, clinically apparent liver disease, bloody diarrhea), if the pregnancy was abnormal, or if they had already enrolled during a previous pregnancy. The mothers and infants underwent intensive surveillance during the first year of infant life. Blood samples were taken and stored from both mother and cord blood around the time of birth. Samples, including whole blood, were then obtained from the child annually.^39^. All infants under follow up had a sample of whole blood collected annually on or around their birthday (2-5 ml depending on the age). The child’s samples were subsequently divided into plasma and red cell pellets as described in more detail below. Infants were included in the present study if 1) receipt of three doses of DTwP/Hib/HBV (at approximately 6, 10 and 14 weeks of age) and one dose of MV vaccine (at 9 months of age) could be confirmed as being administered by the research team or from their vaccination records 2) DNA could be extracted from stored red cell pellets 3) plasma samples were available from the 12 month age point of sampling. Informed written consent was re-acquired from the mothers or guardians, and where appropriate consent from the child and assent from the guardian or mother, specifically for the genetic component of this study. Ethical approval was provided locally by the Uganda Virus Research Institute (reference GC/127/12/07/32) and Uganda National Council for Science and Technology (MV625), and in the UK by London School of Hygiene and Tropical Medicine (A340) and Oxford Tropical Research (39-12 and 42-14) Ethics Committees.

##### South Africa: The Soweto Vaccine Response Study

Six-month infants born in Chris Hani Baragwanath Hospital living in the Soweto region of Johannesburg, South Africa were identified from screening logs and databases of participants involved in vaccine clinical trials^40^ coordinated by the Vaccine and Infectious Diseases Analytics (Wits-VIDA) Unit (https://wits-vida.org). Mothers of the infants were approached if the infants had received all of their vaccines up to six months of age (DTaP/Hib/HBV at approximately 4, 8 and 12 weeks of age). After receiving information about the study the mothers were consented in accordance with ethical approval from the University of Witwatersrand Human Research Ethics Committee (reference M130714) and the Oxford Tropical Research Ethics Committee (1042-13 and 42-14). The infants were sampled prospectively at six months of age and at 12 months after receipt of MV vaccine at 9 months. Single whole blood samples were collected and prepared using a similar protocol to that used in Entebbe to extract DNA from cell pellets and plasma for antibody assays.

##### Burkina Faso: The VAC050 ME-TRAP Malaria Vaccine Trial

Infants between the ages of 6 and 18 months living in the Banfora region of Burkina Faso were recruited into a Phase 1/2b clinical trial to test the safety, immunogenicity and efficacy of an experimental heterologous viral-vectored prime-boost liver-stage malaria vaccine ^41^. These infants were all expected to receive their EPI vaccines (DTwP/Hib/HBV) as part of the usual national schedule at 4, 8 and 12 weeks of age. Infants were precluded from participating in the trial if they were found to have clinical or hematological (venous hemoglobin less than 8 g/dL) evidence of severe anemia, history of allergic or neurological disease or malnutrition. Of a total of 730 infants that were recruited into the study following informed and written consent from the mother, samples suitable for extraction of DNA were collected and stored from 400 infants (350 vaccine recipients and 50 recipients of a control rabies vaccine). Samples of plasma were available from the infants at multiple time-points following the experimental vaccine receipt. Samples from individuals taken at time points as close to the 12-month age as possible were prioritized for EPI vaccine response measurements. The infants underwent intensive clinical history and examination during screening and follow-up. The mothers of the participating infants provided consent for their children to be enrolled in the clinical trial and for subsequent genetic studies to be undertaken for all vaccines received in accordance with ethical approval from the Ministere de la Recherche Scientifique et de l’Innovation in Burkina Faso (reference 2014-12-151) and the Oxford Tropical Research Ethics Committee (41-12).

#### Avon Longitudinal Study of Parents and Children

Genotype data was available from ALSPAC as described previously ^24, 42, 43^ and selected using the fully searchable data dictionary and variable search tool (http://www.bristol.ac.uk/alspac/researchers/our-data/). Consent for biological samples was collected in accordance with the Human Tissue Act (2004) and ethical approval for the study was obtained from the ALSPAC Ethics and Law Committee and the Local Research Ethics Committees.

### Laboratory methods

#### 1000Gp3 and HapMap DNA extraction

Commercially available plates of DNA extracted from LCLs (ACB: MGP00016; ASW: MGP00015; ESN: MGP00023; GWD: MGP00019; LWK: MGP00008; MSL: MGP00021; YRI: MGP00013) and individual aliquots of DNA from cell lines of MKK samples (**Table S4**) were all acquired from Coriell Institute for Medical Research (New Jersey, USA).

#### VaccGene blood sampling and preparation

Whole blood was sampled into vacutainer tubes (BD, Becton Dickinson and Company, New Jersey, USA) containing ethylenediaminetetraacetic acid (for the Ugandan and South African studies) or lithium heparin (Burkinabe) as an anticoagulant. Following centrifugation the samples were separated into their constituent parts (plasma, buffy coat and red cell / erythrocyte layers) and stored at −80°C until downstream analysis in batches. DNA was extracted from the erythrocyte layer in the Ugandan study and from the buffy coat in South African and Burkinabe studies. DNA from all cohorts was extracted from the relevant samples using Qiagen QIAamp DNA Mini or Midi Kits (Qiagen, Hilden, Germany) using recommended protocols. Whole blood was also sampled into serum separator tubes (SST; BD, New Jersey USA) in the Ugandan study and serum was isolated and stored according to the recommended protocols.

#### HLA classical allele typing

6- digit ‘G’ resolution HLA typing was performed for all African samples using a commercial platform developed by Histogenetics (Ossining, New York, USA). Whole gene long-read sequencing was performed using PacBio technology for a subset of African individuals and loci. A more detailed description of exons typed and nomenclature can be found in the **Supplementary Text**. Exon targeted MiSeq (Illumina, California, USA) sequencing was performed by Histogenetics (Ossining, New York, USA) following preparation of libraries from individual DNA according to MiSeq protocols with two amplification rounds tagging adaptor and index sequences followed by sequencing on a MiSeq machine according to manufacturer protocols. The resultant fastq files were processed and typed using proprietary HistoS and HistoTyper softwares (Histogenetics, New York, USA) ^44^ using IMGT/HLA Release 3.25.0 July 2016. Gene-targeted PacBio sequencing was undertaken by HistoGenetics on the RS II using standard protocols with a FastQ file produced from the SmartAnalysis pipeline. Subsequent typing results were generated using the proprietary HistoS and HistoTyper reporting softwares ^44^. Sequence reads achieved a depth of at least 100x coverage of the targeted exons. A subset of 90 individuals from Uganda were also typed using Sanger-sequence based HLA typing performed by an accredited tissue typing laboratory at Addenbrooke’s Hospital, Cambridge University Hospitals NHS Foundation Trust using the proprietary uTYPE software version 7 (Fisher Scientific. Pittsburgh, USA). The list of possible ambiguous calls were minimized by using the ‘allele pair’ export function in this software which lists all possible and permissible allele pair possibilities for each locus for each individual. Alleles were defined using the IMGT/HLA Release: 3.22.0 October 2015. Best-call allele pairs for each locus in each individual were determined based on local guidelines prioritizing alleles that were ‘Common and Well-Documented’ (CWD) ^45^ but any genotype inconsistencies were highlighted and inspected manually for potential evidence of novel mutation. In a subset of the 1000Gp3 populations, allele calls were available from a previous round of lower resolution (4-digit or 2-field) typing using Sanger sequencing ^46^. These calls were used to test reliability of typing and estimate reductions in ambiguity calls for African, CHS and GBR individuals.

#### Quantitative vaccine response antibody assays

Three validated multiplex immunoassays were used to measure antibody concentrations against a number of vaccine antigens in the three *VaccGene* populations. Briefly, this method measures total IgG against each respective antigen including functional (e.g. neutralizing) as well as non-functional antibodies. Antibodies against DT, TT, pertussis toxin (PT), pertactin (PRN), filamentous haemagglutinin (FHA), and MV were determined in the MDTaP assay which is a combination of two previously described assays^47, 48^. Antibodies against Hib polysaccharide were determined in the HiB assay^49^. For MV and DT the correlation of the multiplex immunoassay to gold standard functional assays is high^50, 51^. The immunoassay uses Luminex technology (Luminex Corporation, Austin, Texas, USA) that depends on conjugation of commercially available or in-house developed antigens to fluorescent carboxylated beads using a two-step carbo-diimide reaction to covalently link each antigen to a uniquely fluorescing bead. For the MDTaP assay, serum samples were diluted 1/200 and 1/4000 in phosphate buffered saline (PBS)/Tween-20/3% bovine serum albumin and incubated with the beads to allow the binding of any antibody present in the medium whilst minimizing background in a manner similar to a monoplex solid-phase enzyme-linked immunosorbent assay (ELISA). The bead-antigen-antibody complexes were then separated from remaining plasma or serum through the use of a vacuum manifold before washing with PBS and incubating with a further anti-human IgG antibody conjugated to R-phycoerythrin (R-PE), and washing again prior to detection in the Luminex flow cytometer. The HiB assay was performed similarly, with the exception that samples were diluted 1/100 in 50% antibody depleted human serum (ADHS). The cytometer was used to firstly detect the identity of the fluorescently labelled bead (and therefore antigen bound), and then secondly to detect the fluorescence intensity of R-PE (related to the concentration of primary antibody in solution) bound to each bead passing through the detection channel^48^. The final concentration of bound antibody was calculated by determining the median fluorescence intensity of the antigen-specific beads and using diluted standards to calculate the concentration in international units for each antigen. ELISA results were available for MV vaccine and TT antibody responses from a subset of the Entebbe participants as performed as part of the early investigation undertaken in the Ugandan cohort^38^. Hepatitis B surface antigen (HBsAg) responses were measured using the anti-HBs kit on the ABBOTT Architect i2000 using recommended protocols (Abbott Laboratories, Chicago IL, USA).

#### Genome-wide genotyping

SNP Genotyping was undertaken for the three *VaccGene* populations using the Illumina HumanOmni 2.5M-8 (‘octo’) BeadChip array version 1.1 (Illumina Inc., San Diego, USA), performed by the Genotyping Core facilities at the Wellcome Sanger Institute (WSI). Genomic DNA underwent whole genome amplification and fragmentation before hybridization to locus specific oligonucleotides bound to 3μm diameter silica beads. Fragments were extended by single base extension to interrogate the variant by incorporating a labelled nucleotide enabling a two-color detection (Illumina, 2013). Genotypes were called from intensities using two clustering algorithms (Illuminus and GenCall) in GenomeStudio (Illumina Inc., San Diego, USA) incorporating data from proprietary pre-determined genotypes.

#### Whole-genome sequencing of MKK

Whole-genome sequencing to a 30x coverage was undertaken for the MKK using the Illumina HiSeq X platform using a PCRfree library preparation with a PhiX control spike-in on a barcoded tag. Basecalling was performed on the instrument by using Illumina’s sequencing control software (SCS version 3.3.76) and the realtime analysis (RTA) software. The resulting basecalls were converted directly to unmapped BAM format using the WSI’s BAMBI software (version 0.9.4) for injection into our mapping pipeline. The mapping pipeline first removes any adaptor sequence from the SEQ portion of the read and annotates it as an AUX tag to be replaced in the SEQ after mapping as a soft clipped sequence. A spatial filter was next generated for the lane to remove any bubble induced artefacts from the flowcell by mapping the Phi-X sequence to the reference using BWA MEM (version 0.7.15-r1140) and using this to create a mask to remove any contiguous blocks of spatially oriented INDELs using our spatial filter program (pb_calibration version 10.27) after alignment. Meanwhile the human data was mapped to HS38dh using BWA MEM (version 0.7.15-r1140). The output from this process was then converted from SAM to BAM using scramble (version 1.14.8); headers were corrected using samtools reheader (version 1.3.1-npg-Sep2016); and then the data was sorted and had duplicates marked using biobambam (version 2.0.65). Any stray PhiX reads were removed using AlignmentFilter (version 1.19) and the resulting CRAM file was delivered to our core IRODS facility for storage and transfer to the EGA.

Single sample variant calling to GVCF format was performed using GATK HaplotypeCaller (version 3.8-0-ge9d806836). GVCFs were combined into a single GVCF using GATK CombineGVCFs (version 2017-11-07-g45c474f) and then the final VCF callset was created using GATK GenotypeGVCFs and genomic coordinates lifted over to build 37 using LiftOver.

#### RNA sequencing of 1000Gp3 lymphoblastoid cell lines

A custom RNA-Seq read alignment approach was used to identify expression quantitative trait loci (eQTLs) for the *HLA* genes. The HLA region presents a major challenge in determining RNA-Seq based gene expression quantification due to the abundance of paralog sequences that are highly polymorphic. We therefore aligned the short RNA-Seq reads to a reference sequence defined per individual, complemented with alternative HLA alleles in order to improve the mapping of the reads. The eQTL analysis involved the quantification of expression of the following 9 *HLA* genes: HLA-A, HLA-B, HLA-C, HLA-DQA1, HLA-DQB1, HLA-DPA1, HLA-DPB1, HLA-DRB1 and HLA-DRB5.

RNA sequencing was undertaken using existing LCLs from 600 unrelated samples from five African populations in the 1000 Genomes Project, including the 97 LWK, 84 MSL, 112 GWD, 99 ESN, 42 YRI from 1000Gp3 as well as 166 MKK from the HapMap project. Cell lines were retrieved from Coriell in pre-assigned batches. In order to reduce batch effects the samples were divided into batches for sequencing representative of all six populations. Cell cultures were expanded and 1×10^7^ cells/line were pelleted, treated with RNAProtect (Qiagen) and stored at −80 °C until shipment. Following further randomization, RNA extraction from the entire pellets was performed by Hologic/Tepnel Pharma Services using the RNeasy PLUS mini kit (Qiagen). Library preparation was then performed using the standard automated Kapa stranded mRNA library preparation protocol, followed by RNA sequencing on the HiSeq 2500 using paired end sequencing with 75bp reads. The sequencing was carried out at the Wellcome Sanger Institute where 12 samples, randomised across populations, Coriell batches and Hologic RNA extraction batches were sequenced over two lanes to ensure adequate coverage to quantify gene expression whilst minimising systematic bias.

#### Follicular helper T-cell assay

An Antigen Inducible Marker (AIM) method was used to measure and compare proportions of circulating antigen-specific T_FH_ cells in the circulating blood of donors defined by HLA-DRB1 allele carriage. The AIM assay uses flow-cytometry to detect proportions of antigen-specific follicular helper T (T_FH_) cells defined as co-expressing CD25, OX40 and CXCR5 markers following *ex-vivo* antigen stimulation of PBMC^25^. Based on HLA-DRB1 allele type, 1×10^6^ PBMCs were selected from stored samples collected from consenting participants recruited into studies coordinated by the laboratory of Professor Alessandro Sette investigating immunodominant peptides associated with responses against pertussis ^52^, tuberculosis ^53^, dengue ^54^, and IgE allergy ^55^. The samples were thawed and cultured with 30μg/ml PT (Reagent proteins, USA), 5μg/ml DT (Reagent proteins, USA), 5μg/ml TT (List Biological Laboratories Inc., Campbell, CA), 10 μg/ml phytohaemagglutinin (PHA, Sigma, St Louis, MO, USA), or toxoid diluent (water) at 37°C for 24 hours. The cells were then washed, labelled with an antibody panel for 15 minutes at 4°C before being fixed with paraformaldehyde (Sigma, St Louis, MO, USA) and acquired on an LSRII (Becton, Dickinson and Company, New Jersey, USA). The antibody panel was as follows: CCR7-PerCP-Cy5.5 (G043H7), OX40-PE-Cy7 (BerACT35), CXCR5-Brilliant Violet 605 (J252D4) all from Biolegend, San Diego, USA; CD45RA-eFluor450 (HI100), CD4-APC-eFluor780 (RPA-T4) from eBioscience, San Diego, USA; CD25-FITC (M-A251), CD14-V500 (M5E2), CD19-V500 (HIB19), CD8-V500 (RPA-T8) from BD Biosciences, San Jose, USA; LIVE/DEAD Aqua stain (Thermo-Fisher Scientific, Waltham, USA). Data derived from the gating strategy was analysed using FlowJo Software version 10 (FlowJo LLC, Oregon, USA) and either one-tailed Wilcoxon rank sum or linear regression statistical tests were performed in R. All participating donors were known either to have received DT and TT, and either whole cell (wP together known as DTwP) or acellular pertussis (aP, together as DTaP) as part of a vaccine study undertaken in the Sette lab, or self-reported having received standard vaccines during childhood.

#### Cell-specific HLA-wide eQTL analyses

HLA typing was performed on DNA extracted from the Database of Immune Cell eQTLs (DICE) dataset^56^ using the same Histogenetics MiSeq protocol described above.

### Analytical methods

#### SNP quality control (QC)

SNP QC was performed separately for each genotyped *VaccGene* cohort using identical steps and using SNPs mapped to Human Genome Build 37. Low quality variants that mapped to multiple regions within the human genome or did not map to any region were removed. Samples with a call rate of less than 97% and heterozygosity greater than 3 standard deviations around the mean were filtered sequentially. Sex check was performed in PLINK (v1.7) using default F values of <0.2 for males and >0.8 for females^57^. Samples with discordance between reported and genetic sex were removed. Genetic variant filtering was performed across the remaining samples and sites called in <97% samples were removed from each population. Identity-by-descent (IBD) was measured within each population. Only samples with IBD >0.9 not known to be twins were removed using a custom algorithm that removed the sample from the pair with the lower variant call rate. Sites in Hardy Weinberg disequilbrium (*P*<10^-8^) were also excluded from future analysis in all individuals, calculated using individuals with IBD <0.05 (hereafter designated ‘founders’). Following the above quality control steps, principal component analysis (PCA) was performed in EIGENSOFT v4.2^58^ for each population and combined with populations representative of other parts of Africa (the ‘AGV dataset’^20, 59^) or global populations including 1000 Genomes^60^ (‘Global + AGV dataset’). PCA was carried out after LD pruning to a threshold of r2=0.5 using a sliding window approach with a window size of 50 SNPs sliding 5 SNPs sequentially. Regions of long range LD were removed from the analysis. Individuals with values of the first 10 principal components more than six standard deviations around the mean of other samples in each population were removed.

#### Genotype imputation

Haplotype phasing was undertaken in each *VaccGene* population separately using SHAPEIT2^61, 62^ with standard parameters and the advised effective population size of 17,469. We subsequently used IMPUTE2 to estimate unobserved genotypes using a combined reference panel consisting of the 1000Gp3 reference panel^60^ combined with data from the African Genomes Variation Project^20^ and a 4x whole genome sequence coverage dataset of another Ugandan population of 2000 individuals entitled the UG2G dataset: 1000G/AGVP/UG2G^20^.

#### Cohort genotype variant merging

A high quality set of autosomal genotype calls free of batch effects were required for a number of downstream analyses. Variant calls derived from a combination of array genotyping (Illumina omni2.5M passing QC in the *VaccGene* and some 1000Gp3 cohorts) and next-generation sequencing (NGS) for other 1000Gp3 populations (using only calls at sites intersecting with omni2.5M typed locations) were defined. A comparison of variant calls between array and NGS platforms was undertaken for a subset of 1000Gp3 individuals who had data from both platforms using concordance. Only those sites with concordance estimates of r^2^>0.99 were taken forwards for further analyses. Variants typed on the omni2.5M array were called in all individuals using array genotypes as first priority (where data was available from both array and NGS platforms) and then using NGS data (if array data was not available). Once variant calls were available for all individuals, these variants were used to calculate principal components and ADMIXTURE analysis across all autosomes to ensure that there was minimal evidence of batch variation caused by a differential use of NGS or array variants across individuals and populations.

#### Measuring differentiation of HLA alleles across African and global populations

G_ST_ was calculated for each locus using alleles described in 2-, 4- and 6-digit resolution using the ‘diveRsity’ package in R^63^. G_ST_ and Jost’s *D* statistic^64^ are statistics explicitly designed for multi-allelic residues. Both statistics were calculated but given the close correlation between the two outputs, the availability of G_ST_ statistics in other studies of HLA in Africa^65^ made this the statistic of choice. Allelic richness was calculated in diveRsity using bootstrap sampling (1000 samples) with replacement to estimate the average number of alleles observed with standard errors given the differing number of individuals observed in each population and the likelihood of observing rare alleles.

#### Vaccine antibody response normalization

Measured antibody responses were normalized using both logarithmic and inverse normalization (INT) in R version 3.5.1. Inverse normalized traits were tested for association with a variety of available metadata endpoints to determine covariates to include in the final regression model to increase power in the quantitative analysis^66^. Endpoints included time between vaccination and sampling, sex, age, weight-for-length z score at birth, number of illnesses, socio-economic status and HIV status (if known). Only time between vaccination and sampling was used in the final models. INT trait measures were used throughout our analyses and all results reported as such.

#### Intra-cohort genotype association testing and meta-analysis

Multiple software packages are available that can account for population structure and cryptic relatedness in genomic association studies through the use of mixed model approaches ^67^. However, until recently only a handful of these algorithms could simultaneously account for probabilities of imputation accuracy in large datasets. We therefore applied a mixed model in our association analyses implemented in the GEMMA software^68^ that explicitly accounts for imputed genotypes. We calculated the relatedness matrices using only those autosomal variants directly typed in each population. Inclusion of the first 10 principal components did not affect the association statistics for any tested phenotype in any cohort as would be expected given that these models explicitly account for population structure and relatedness and so these PCs were not included in any downstream association testing. The METASOFT software was used to undertake fixed and random effect meta-analysis to test for shared signals of association across populations^69^.

#### HLA imputation and HLA reference panel construction

The HLA*IMP:02 software was used for imputing classical HLA alleles to 2- and 4-digit resolution at all 11 loci in *VaccGene* individuals with available genotype data^22^. HLA*IMP:02 was used preferentially above other software including SNP2HLA^70^ and HIBAG^71^ because of 1) the inclusion of individuals of West African ancestry in the reference panel of HLA*IMP:02 and reported accuracies of imputation of individuals from diverse population backgrounds^22^, 2) the explicit handling of missingness of types between individuals and 3) the adaptability of the algorithm by our team to allow for higher resolution types and amino acid imputation. Imputation of HLA alleles in the African and UK (ALSPAC) populations was performed a) using the March 2016 release of the HLA*IMP:02 reference panel using default settings to establish a baseline for accuracy and b) using an African-specific reference panel with algorithmic modifications, described below. The ‘best-guess’ call was defined for each diploid allele in every individual using the output from the algorithm in the presence or absence of an imposed threshold for calling using the posterior probability of 0.7. It has been proposed that imposing this threshold improves the quality of the total number of calls at the expense of reducing the total number of available calls. In downstream association analyses, this posterior probability was used as variant dosages to account for probabilities in regression analyses.

The African-specific reference panel was built using only variants (derived from publically available array genotype or whole-genome sequence data for 1000Gp3 and MKK populations or array genotypes for the *VaccGene* populations as described above) and 6-digit ‘G’ calls from the 1,705 typed individuals. Five-fold cross validation, comprising five random splits of the reference dataset into training (four-fifths of the data) and validation (one-fifth of the data) sets, was carried out to evaluate expected imputation accuracy on African samples. For each split, accuracy in the validation set was assessed using the metrics described below. All imputations used for association analyses were based on the complete reference panel.

Comparisons between imputed vs typed calls were undertaken at the 4-digit (i.e. 2-field) level of resolution. If an available call at a single allele locus included several potential higher resolution alleles (i.e. a list of potential ambiguities) only the first available allele call from either platform (adhering to a CWD priority) were used for comparison. In the cases of comparing imputed HLA calls to typed calls, any 6-digit ‘G’ type calls were reduced to 4-digit and treated as the ‘truth’ set. By comparing each individual allele in turn it was possible to define calls of the test platform that were:

- True positives (*TP*)
- False positives (*FP*); called by the test platform as that allele when it was in fact another allele according to the truth)
- False negatives (*FN*; called by the test platform as another allele when it was in fact this allele)
- True negatives (*TN*).

Thus at the level of an individual allele various metrics could be calculated. Sensitivity was defined as:

*TP / (TP + FN)*

Specificity was defined as:

*TN / (TN + FP)*

Positive predictive value (PPV) was defined as:

*TP / (TP + FP)*

Negative predictive value (NPV) was defined as:

*TN / (TN + FN)*

Accuracy was defined as:

*(TP + TN) / (TP + FP + FN + TN)*

Concordance was calculated at the level of the locus. For every pair of chromosomes with data available in both truth and test sets the number of identical allele calls between platforms was calculated and divided by the total number of alleles, equivalent to the positive predictive value (PPV). Any individual with missing alleles on either or both chromosomes on either platform were excluded from these calculations.

HLA imputation using the Broad Multi-Ethnic panel was performed using the Multi-Ethnic HLA reference panel (version 1.0 2021) available on the Michigan imputation server using recommended settings^23^.

#### Pooled linear mixed model and HLA variant association testing

In order to undertake conditional analyses including all genotyped and imputed genotype variants across the HLA locus in addition to HLA allele and amino acid variants across all three populations we leveraged the intra-cohort normalized, quantitative nature of the antibody responses and combined all individual level genetic data from individuals in all three *VaccGene* populations maintaining imputation dosages where appropriate. For HLA alleles and amino acids, posterior probabilities were used to infer imputation dosages at each allele. We calculated a relatedness matrix using only directly genotyped autosomal variants from the three populations and we then undertook association testing using dosages in GEMMA to account for imputation probabilities in the context of both imputed genotypes and HLA alleles and amino acid variants. The resultant *P*-value association statistics were then compared to output from the fixed effects meta-analysis approach determined using METASOFT using the Pearson correlation coefficient. Step-wise forward conditional modelling was used for each trait including the index SNP dosages as fixed effect covariates in the model to assess for evidence of interdependence whilst taking differential LD patterns into account across all populations.

#### Fine-mapping HLA associations with each trait

An approach similar to that used by Moutsianas and colleagues investigating the effect of HLA in multiple sclerosis^72^ was used to compare and contrast the results of both manual and automated step-wise linear modelling approaches. First, stepwise conditional modelling was performed using the pLMM approach in GEMMA for each trait to identify independently associated loci achieving a significance threshold of *P*≤5×10^-9^. This approach resulted in a range of SNPs, HLA alleles or amino acids likely to be independently associated with each trait, frequently spanning multiple loci across the class II region. The gene origins of these ‘independent index’ variants were determined (SNP or amino acid residues in HLA-DRB1 for example) and the dosages of all variants were then incorporated in a manual modelling approach. For this manual approach, a refined number of unrelated individuals (IBD<0.2) were selected and models of association were tested using additive dosage probabilities for imputed genotype, classical allele and bi-allelic amino acid residues across all 11 loci with a population average minor allele frequency (MAF_AV_) greater than 0.01. Null models were defined for each trait by including the first five genetic principal components and the ‘time between sampling most recent vaccination’ covariate. Independent index variants discovered through the pLMM analyses were assessed both in *univariate* (i.e. single SNP, HLA allele or bi-allelic amino acid residue variable) models or *multivariable* (i.e. defining more than one single SNP, HLA allele or amino acid residue) models. Models were rationally tested and compared based on the known associations between amino acid residues and classical alleles. For example, an arginine at position 74 in the HLA-DRB1 protein (designated DRB1-74Arg) is only found in alleles in the 2-digit HLA-DRB1*03 allele group. Using the 6-digit ‘G’ resolution the only allele groups therefore containing DRB1-74Arg include HLA-DRB1*03:02:01 and HLA-DRB1*03:01:01G. Each model defined using this framework was tested and compared. Using the given example, univariate models comparing the DRB1-74Arg and HLA-DRB1*03 variants, and a conditional model including both HLA-DRB1*03:02:01 and HLA-DRB1*03:01:01G would be compared. All models included the same principal components and time covariates as defined in the null model for each trait. The models were compared to the null using the likelihood ratio test (LRT) if the models were nested, or using the Bayesian Information Criterion (BIC) otherwise. Models with lower BIC values were interpreted to explain the variance in the observed data most parsimoniously.

Finally, any prior knowledge from the associations derived from the LMM associations were removed and automated bidirectional stepwise model selection based on the BIC was undertaken. This modelling was designed to test whether models incorporating amino acid residues or classical alleles best explained each trait at each locus and also to determine whether any other variants should be considered in a final model other than those identified using the manual approach above. A consensus model was then determined based on the results of the manual and automated approaches for each trait. Manual and automated modelling steps were performed in R 3.5.1.

Given the relatively small size of the dataset compared to existing efforts for other diseases including multiple sclerosis^17^ and inflammatory bowel disease^18^ only additive models of association were tested. Deviation from additivity or interaction between HLA variants was not assessed because our study was likely to have insufficient power to detect such effects.

#### RNA Sequencing and eQTL Analysis

RNA sequencing reads were inspected using the FastQC tool for quality control. Reads were trimmed using Cutadapt for polyA and adaptors prior to mapping. The merged set of whole-genome genotypes derived from a combination of array and sequencing data from VaccGene, 1000Gp3 and Hapmap samples was used for the eQTL data analysis. All samples with RNA-Seq data available also had genotype data available. Variant calls from both genotype and sequence data for these samples were included in eQTL analyses. After accounting for QC of the RNA sequence data, there was a total of 558 samples available for the eQTL analysis: ESN (99), GWD (112), LWK (97), MKK (126), MSL (83), and YRI (41).

The RNA-Seq data set was mapped to a custom genome reference sequence that consisted of the non-HLA containing human reference sequence (hg38) and HLA containing reference sequence unique to each individual. The HLA-containing reference was generated based on the 6-digit ‘G’ type results of the samples in our dataset. We extracted a total of 285 HLA alleles: 47 HLA-A, 73 HLA-B, 35 HLA-C, 11 HLA-DPA1, 39 HLA-DPB1, 8 HLA-DQA1, 25 HLA-DQB1, 45 HLA-DRB1 and 2 DRB5 nucleotide sequences of exons from the international ImMunoGeneTics/HLA database v3.33.0 at the European Bioinformatics Institute. For each HLA allele, we generated a sequence where the exons of the respective allele were merged with 200 bases of spacers (N) as introns. The exons that were not typed in the ImMunoGeneTics/HLA database for each HLA allele were filled using the closest allele. The resulting HLA containing reference contained 285 HLA gene structures with the corresponding exons and the introns of N characters. We generated an annotation file for the HLA-containing reference in the form of a GTF file as well as the exon-exon junction file for the mapping. Non-HLA containing reference was generated from the human reference sequence (hg38) excluding the alternative haplotype contigs where the 9 HLA genes in the reference were removed from the reference sequence by hard masking. We used the corresponding Ensemble gene annotation (v83) for the Non-HLA reference sequence. The custom reference sequence for the RNA-Seq data mapping was generated by merging the non-HLA containing reference sequences with the HLA containing reference sequences. The annotations and the exon-exon junctions were merged to generate the final gene annotation GTF file for the mapping.

Alignment was performed using the STAR alignment tool ^74^ in two-pass mode. Our custom reference sequence and the custom gene annotations were used for the indexing of the reference sequence for the mapping. During the second pass we used the novel exon-exon junctions as well as the exon-exon junctions we generated for the HLA containing reference. The quantification of RNA transcripts was strongly affected by reads that mapped to multiple locations in the custom reference sequence. Since we had 285 HLA alleles with high similarity in our reference and the default maximum number of multiple alignments in STAR aligner is 10 we increased the maximum number of multiple alignments to 300 for the RNA-Seq mapping. We counted the number of reads mapping to the HLA haplotypes using a custom method using the htslib for accessing the alignment files in bam format. We used two criteria to count the reads: 1) If the reads were mapped to the multiple HLA haplotypes, but no other regions in the genome, we counted these reads as single mapping, 2) If the reads were mapped to a unique HLA allele, the reads were counted for that allele. After verifying the reads were mapping to their correctly typed HLA alleles, we quantified the gene expression for each HLA gene as the sum of these counts. The read counts for the other genes were calculated with htseq-count v0.9.1, using the gene annotations from Ensembl as the features. The counts were merged to include the whole set of gene counts. Normalization was performed using the DESeq2 tool with the variance stabilized transformation^75^. The variance-stabilized transformation was performed after the library size and dispersion estimation. Normalization was performed for each population separately.

eQTL mapping was performed for the 5Mb region that included the nine HLA genes of interest. We restricted our search to cis-eQTLs by selecting variants within 1Mb of each gene’s start and end positions. Per population, cis-eQTLs were identified by linear regression where normalized gene expression was regressed on variant dosage correcting for covariates using Matrix eQTL ^76^. Covariates included population principal components calculated from genotype data, meta-data on known technical variables and unobserved confounding variables detected using Surrogate Variable Analysis (SVA). Per population for each variant we calculated the *P*-values that are corrected using the Benjamini-Hochberg procedure and the beta values. The results of the eQTL analysis for six populations were then combined using a fixed effects model implemented by METASOFT.

The same methods were used for the individual cell types using the DICE dataset. This dataset included 14 cell types in which the effect of a single variant (rs545690952) was explored. The overall significance of association with each cell type was as follows: naïve B-cells (*P*=0.19), naïve CD4 T-cells (0.59), stimulated CD4 T-cells (0.36), naïve CD8 T-cells (0.99), stimulated CD8 T-cells (0.53), monocytes (8.6×10^-3^), natural killer cells (0.19), T_FH_ (0.27), Th1 (0.86), Th2 (0.68), Th17 (0.07), Th* (0.42), Tregmem (0.83), Tregnaive (0.56).

To test the reproducibility of our approach, we replicated a well-characterized eQTL for HLA-C associated with differential control of HIV-1^77^ in the 1000Gp3 dataset. We observed a strong effect of rs2395471 on HLA-C expression in the African populations (*P*=1.14×10^-12^) in the same direction as reported previously.

#### Trait and genetic correlation

Correlation between normally distributed continuous variables or traits were tested using Pearson’s correlation coefficient. Equivalent testing for variables or traits not considered continuous or sufficiently normalized were undertaken using Spearman rank. Testing for the significance of correlation between HLA amino acid residues derived from the present study and a historical GWAS of self-reported pertussis^24^ was performed using permutation. The null distribution was calculated by randomly assigning different SNP identities to the calculated beta coefficients from the pertussis GWAS and recalculating Pearson’s r between 100,000 to 100,000,000 times (dependent on whether a *P*-value could reliably be calculated). The *P*_perm_ value was calculated as the frequency at which a Pearson’s r value calculated from permutation was observed to surpass the r from the true data. These calculations were undertaken using both complete variant datasets and datasets pruned by LD (keeping only the top associated SNP and those SNPs with r^2^<0.35).

#### Peptide binding assays

The Immune Epitope Database (IEDB^78^) was used to test whether the affinity or breadth of peptides derived from specific protein sequences differed by groups of HLA alleles defined as being associated with increased or decreased antibody responses. The output from the binding prediction algorithm included a binding affinity prediction (IC_50_ - measured in nM) and a percentile rank generated by comparing the predicted IC_50_ against scores of 5,000,000 random 15-mers selected from the SWISSPROT database^79^. The percentile rank scores of 15-mer peptides derived from PT (GenBank accession ALH76457), DT (BAL14546) and TT (WP_011100836) were compared. The highest affinity peptide per protein and allele was defined using the peptide with the lowest percentile score. To increase power to identify differences between groups of alleles, all HLA-DRB1 alleles present in the IMGT database were divided into groups dependent on their sequences and whether they possessed an excess of residues associated with either increased (defined as ‘DRB1-233Thr’ alleles for PT) or decreased (defined as ‘DRB1-233Arg’ alleles) antibody responses. The definition of these alleles for PT vaccine responses was undertaken as follows. Firstly the number of residue positions found to be significantly (*P*<0.05) associated with either PT (n=39) responses were determined and then alleles were defined as to whether they had an excess (>1.5x) of residues associated with either a positive beta or those with an excess (>1.5x) of negative beta effect estimates. The distributions of affinities of the top-predicted binding peptides for each of the alleles classified as such were then compared and tested for differences using a two-tailed Mann-Whitney U test. The breadth of antigen-specific peptide binding by class II HLA alleles was defined by measuring the proportion of peptides predicted to bind within the top 5th percentile of all peptides from each peptide per allele of interest, compared across antigens and allele groups.

### Data availability

All direct genotypes from *VaccGene* individuals post-quality control alongside imputed data and raw and curated HLA sequence data and calls have been submitted to the European Genome-Phenome Archive under accession EGAS00001000918. Summary statistics for the genome-wide association tests of imputed data for eight vaccine antibody levels are available on Zonodo (https://doi.org/10.5281/zenodo.7357687).

## Notes

### Competing Interest Statement

The authors have declared no competing interest.

### Funding Statement

This project has received funding from the European Research Council (ERC) under the European Unions [Seventh Framework Programme (FP7/2007-2013)] (grant agreement No 294557).
This work was supported by the Wellcome Trust grant numbers 064693, 079110, 095778, 217065, 202802 and 098051.
AJM was supported by an Oxford University Clinical Academic School Transitional Fellowship and a Wellcome Trust Clinical Research Training Fellowship reference 106289/Z/14/Z and the National Institute for Health Research (NIHR) Oxford Biomedical Research Centre (BRC).
NJT is a Wellcome Trust Investigator (202802/Z/16/Z), is the PI of the Avon Longitudinal Study of Parents and Children (MRC & WT 217065/Z/19/Z), and is supported by the University of Bristol NIHR Biomedical Research Centre (BRC-1215-2001), the MRC Integrative Epidemiology Unit (MC_UU_00011) and works within the CRUK Integrative Cancer Epidemiology Programme (C18281/A19169).
Computation used the BMRC facility, a joint development between the Wellcome Centre for Human Genetics and the Big Data Institute supported by the NIHR Oxford BRC.
Financial support was provided by the Wellcome Trust Core Award Grant Number 203141/Z/16/Z.
Computational support and infrastructure was also provided by the Centre for Information and Media Technology (ZIM) at the University of Dusseldorf (Germany).
The UK Medical Research Council and Wellcome (Grant ref: 217065/Z/19/Z) and the University of Bristol provide core support for ALSPAC.
This publication is the work of the authors and NJT will serve as guarantor for the contents of this paper. GWAS data for ALSPAC was generated by Sample Logistics and Genotyping Facilities at Wellcome Sanger Institute and LabCorp (Laboratory Corporation of America) using support from 23andMe.The cellular immunology studies were supported by National Institute of Health grants U19 AI118626 (AS) and U01 AI141995 (AS/BP).
The views expressed are those of the author(s) and not necessarily those of the NHS, the NIHR or the Department of Health.

### Author Declarations

Uganda Virus Research Institute (reference GC/127/12/07/32) gave ethical approval for this work Uganda National Council for Science and Technology (MV625) gave ethical approval for this work London School of Hygiene and Tropical Medicine (A340) gave ethical approval for this work Oxford Tropical Research Ethics Committees (39-12 and 42-14) gave ethical approval for this work University of Witwatersrand Human Research Ethics Committee (reference M130714) gave ethical approval for this work Oxford Tropical Research Ethics Committee (1042-13 and 42-14) gave ethical approval for this work Ministere de la Recherche Scientifique et de Innovation in Burkina Faso (reference 2014-12-151) gave ethical approval for this work

