## Supplementary Materials for "High-resolution African HLA resource uncovers *HLA-DRB1* expression effects underlying vaccine response"

**This PDF file includes:**

Supplementary Text
Figs. S1 to S3
Tables S1 to S10
Additional Data Tables 1 to 4

### Supplementary Text

<u>HLA allele nomenclature and methods used for typing</u>

Throughout this manuscript, HLA alleles were classified according to the World Health Organization Nomenclature Committee for Factors of the HLA System (*62*). All alleles have an 'HLA' prefix followed by a hyphenated gene name and a subsequent star separator. Two or three digit fields of between two to three digits in length each then follow this prefix separated by colons. The first field (frequently two digits in length in modern nomenclature and therefore referred to as '2-digit' level of resolution) refers to the serological group. The second field (two or three digits long, often referred to as '4-digit' level of resolution) refers to resultant amino acid sequences giving rise to the HLA protein. Alleles that differ in the second field must have at least one identified non-synonymous difference between them. The third field (usually two digits in length) identifies sequences divergent due to synonymous variation (equivalent to '6-digit' resolution). The fourth field refers to variation within the introns ('8-digit' resolution). The allele may be suffixed by a set of letters including an 'N' that donates known 'null' expression of the gene product.

Traditionally, HLA calls have been defined based on variation within exons of the genes that encode the peptide binding domains (PBD; exons 2 and 3 for class I and exon 2 for class II). Therefore, true sequence diversity across all other exons and introns for each gene is relatively unknown, although reference databases are continually accruing extended sequences for many described alleles. As a consequence of this observation, a '6-digit G' level of resolution has been determined whereby alleles can be suffixed by a 'G' donating that the sequence of the exons encoding the peptide binding domain of that gene would be consistent with a 'group' of alleles. These groups of alleles are defined according to a list maintained by the IMGT/HLA working group (http://hla.alleles.org/wmda/hla_nom_g.txt accessed on 20 April 2016).

Although there is substantial data available for worldwide HLA types, many such datasets have been generated using various methodologies spanning sequence specific oligonucleotide and primer technologies through to Sanger and next-generation sequencing (NGS) methods that target variable regions of each gene. A single best allele call is often presented for each chromosome and each individual that often represents a long list of potential ambiguities, and such technologies do not always offer the opportunity to elucidate these ambiguities with challenges in terms of IMGT data releases. With the aid of the increased coverage of exon sequencing possible with the MiSeq platform used for large-scale HLA typing it was possible to reduce both the lists of potential alleles included in 'Groups' and reduce *cis*/*trans* ambiguities through phasing with MiSeq sequencing technology. An amended 'G' list was therefore developed to account for these differences. In the majority of tested individuals it was possible to resolve alleles to a single 6-digit (3-field) call, whereas in some cases a G code was still required. The exons sequenced for all genes using the MiSeq platform included: 2 alone (HLA-DPA1, -DQA1, -DRB3, -DRB4, and -DRB5); 2 and 3 (HLA-DPB1, -DQB1 and –DRB1); 1, 2, 3 and 4 (HLA-A and –B); 1, 2, 3, 4 and 7 (HLA-C).

For a subset of individuals and loci it was possible to undertake near whole-gene PacBio sequencing. Only DNA passing stringent quality and yield thresholds was used for PacBio sequencing and genes were targeted sequentially resulting in sequential attrition of sample availability biased to specific loci. Genes were prioritized in the following order: HLA-B, HLA-A. HLA-C, HLA-DQB1, HLA-DRB1, HLA-DQB1, HLA-DPB1, HLA-DQA1, HLA-DPA1.

47 unrelated individuals from Entebbe were used to validate the MiSeq generated calls by undertaking a comparison of Sanger and MiSeq based typing. The Sanger-based typing method was undertaken using a moderately different set of exon coverage: exon 2 alone (HLA-DRB3, -DRB4, and -DRB5); 2 and 3 (HLA-DPA1, -DQA1, -DQB1 and –DRB1); 2, 3 and 4 (HLA-A and –B, -C and -DPB1). In all cases any observed discrepancies could be resolved by taking into consideration differential exon coverage or the ability to resolve *cis*/*trans* ambiguities using the MiSeq platform. No discrepancies were observed due to differences in IMGT/HLA releases used for calling. The MiSeq platform was deemed superior for large-scale typing owing to the increased ability to resolve ambiguities.

HLA types were available for a subset of the 1000Gp3 samples from an earlier study using an older version of the IMGT/HLA release and using older Sanger sequencing based methods as described in the Methods and Materials. This data was used to demonstrate the utility of our methods compared to traditional methods through reducing ambiguous allele calls (**Fig. S1A**). Using high-coverage WGS data available from the Asian populations of 1000Gp3 and HGDP populations, HLA types were generated across all loci (except DRB3, DRB4 and DRB5) using HLA-LA with cross validation against Sanger and MiSeq based sequencing where available.

A summary of the numbers of African individuals with data generated on each platform (MiSeq, PacBio, Sanger and intersecting array or next-generation sequencing variant calling) is provided in **Table S3**. A summary of the numbers of other populations with HLA allele data available as a reference is provided in **Table S4** and HLA alleles for all individuals are provided in **Additional Data Table 1.**

Novel HLA alleles

The novel alleles described below relate only to exon coding in African populations. These results are summarized in **Fig. S1D** and **Table S5.** Novel alleles discovered in the Oceanian dataset are described in **Table S6**.

*HLA-A*

7 novel alleles were observed in 21 individuals across 7 populations of which 5 were identified using MiSeq (exons 1 and 4) and 2 were only captured using long-read sequencing (including exons 5 and 6).

*HLA-B*

2 novel alleles were observed in 2 individuals (one from ACB and one from Uganda) of which both were detected using MiSeq.

*HLA-C*

5 novel alleles were observed in 5 separate individuals spanning 4 populations. 4 of the novel alleles were detected using MiSeq and 1 using PacBio with novel variation in exon 6.

*HLA-DPA1*
14 novel alleles were identified in 54 individuals from all included populations except Burkina Faso. 10 of the novel were detected using MiSeq and 4 were detected using PacBio with variants in exons 3, 4 and 5.

*HLA-DPB1*
8 novel alleles were identified in 12 individuals from 5 populations of which 5 were identified using MiSeq and 3 were detected with PacBio due to variation in exon 4.

*HLA-DQA1*
7 novel alleles were identified in 16 individuals from 4 populations of which 5 were detected using MiSeq and 2 were identified using PacBio due to variation in exon 4.

*HLA-DQB1*
6 novel alleles were identified in 16 individuals from 6 populations of which 4 were identified using MiSeq and 2 were identified using PacBio due to variation in exon 4.

*HLA-DRB1*
2 novel alleles were observed with one individual in each of ACB and South African populations of which both were detected using MiSeq.

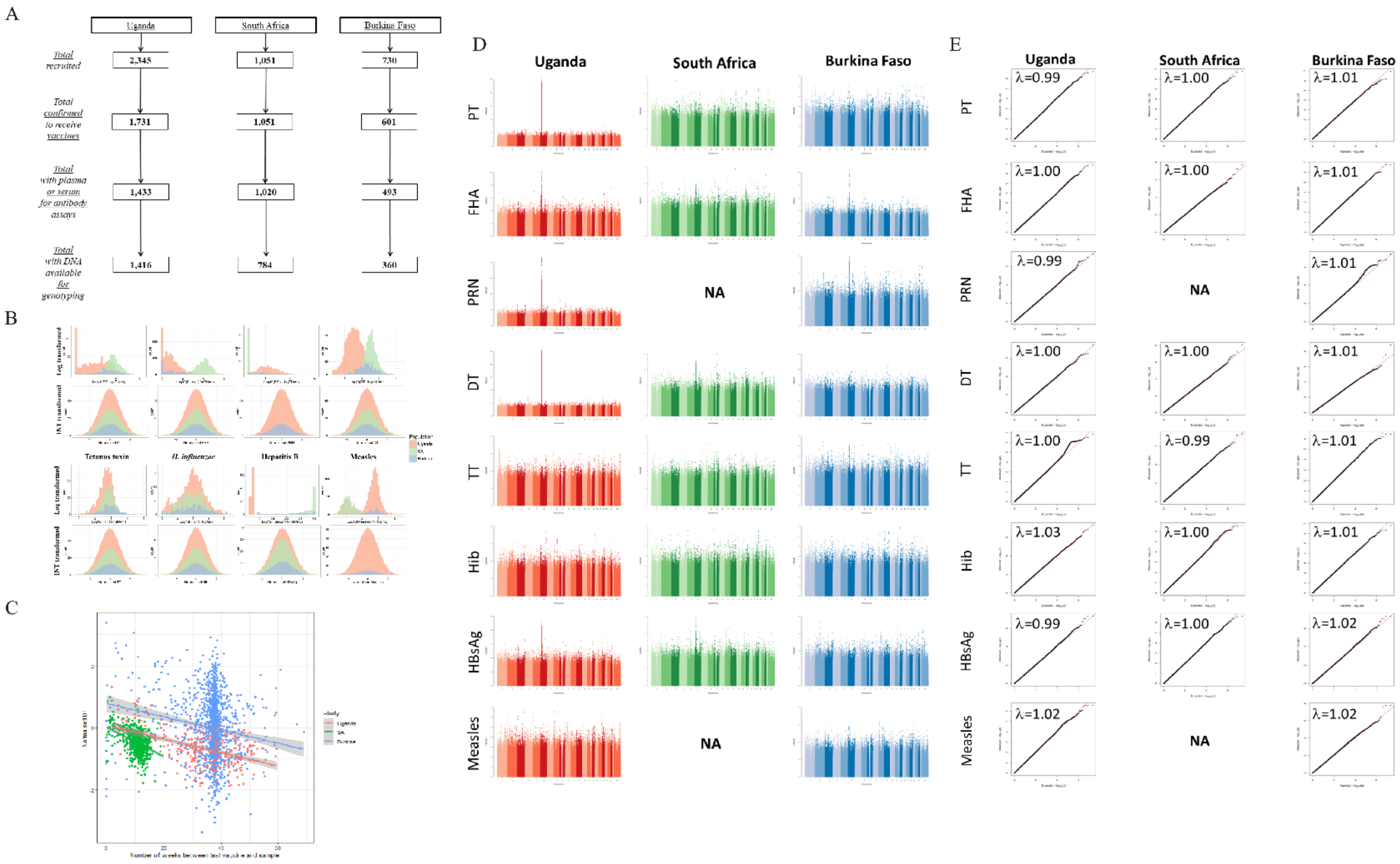


**Fig. S1.**

**A multi-population genome-wide association study of responses to eight vaccine antigens. (A)** Individuals recruited with vaccine-related data and samples available for analysis as part of VaccGene in each cohort. **(B)** Distributions of vaccine responses in individual VaccGene cohorts following transformation using two methods. Distributions of each vaccine response following either logarithmic (log) or inverse normal transformations (INT; used in the GWAS analyses) are shown for each population separated by colour. Individuals in South Africa did not receive pertussis pertactin or measles vaccine prior to the antibody assays being performed.

Differences in log-transformed distributions are due to differences in timing of sampling as shown in **Table S1**. **(C)** Time between final vaccination dose of DT and sampling for measuring response to DT within each population. Linear lines of best fit are shown colored by each population with shaded 95% confidence intervals**. (D)** Manhattan plots of genetic association signals with eight tested vaccine traits in three African populations. **(E)** Quantile-quantile plots of association statistics for all measured vaccine traits in all three populations including all tested variants except those in the extended MHC region (chromosome 6, 25.5-34Mb, build 37).

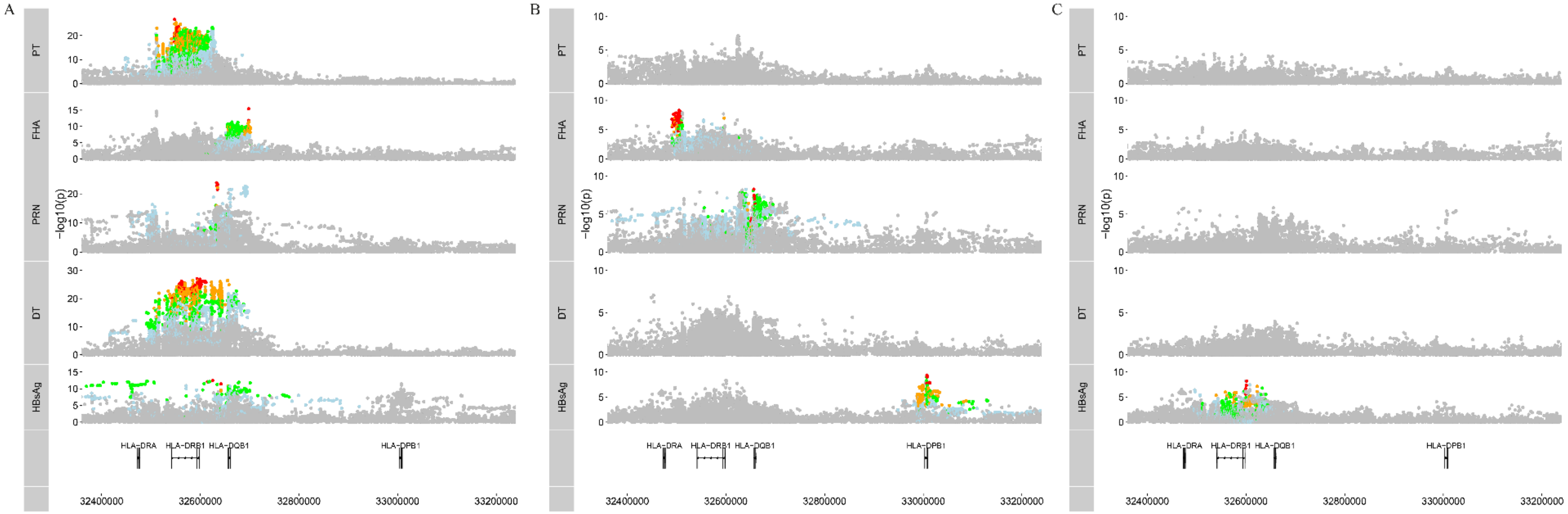


**Fig. S2.**

**Regional plots of association between SNP variants and antibody response traits with and without inclusion of top associated variants as covariates.** Unconditional analyses **(A)**; analyses conditioning on the top associated variant from each individual pooled GWAS **(B)**; and analyses including both variants associated from unconditional and first round of conditional analyses for each individual pooled GWAS **(H)** are shown for each associated vaccine response trait. Wherever the index (most significant *P*-value) variant demonstrated significance $P<5x10^{-9}$, association peaks have SNPs colored by LD ($r^2$) with top associated variant (red 0.8-1; orange 0.6-0.8; green 0.4-0.6; blue 0.2-0.4), otherwise all points are colored in grey.

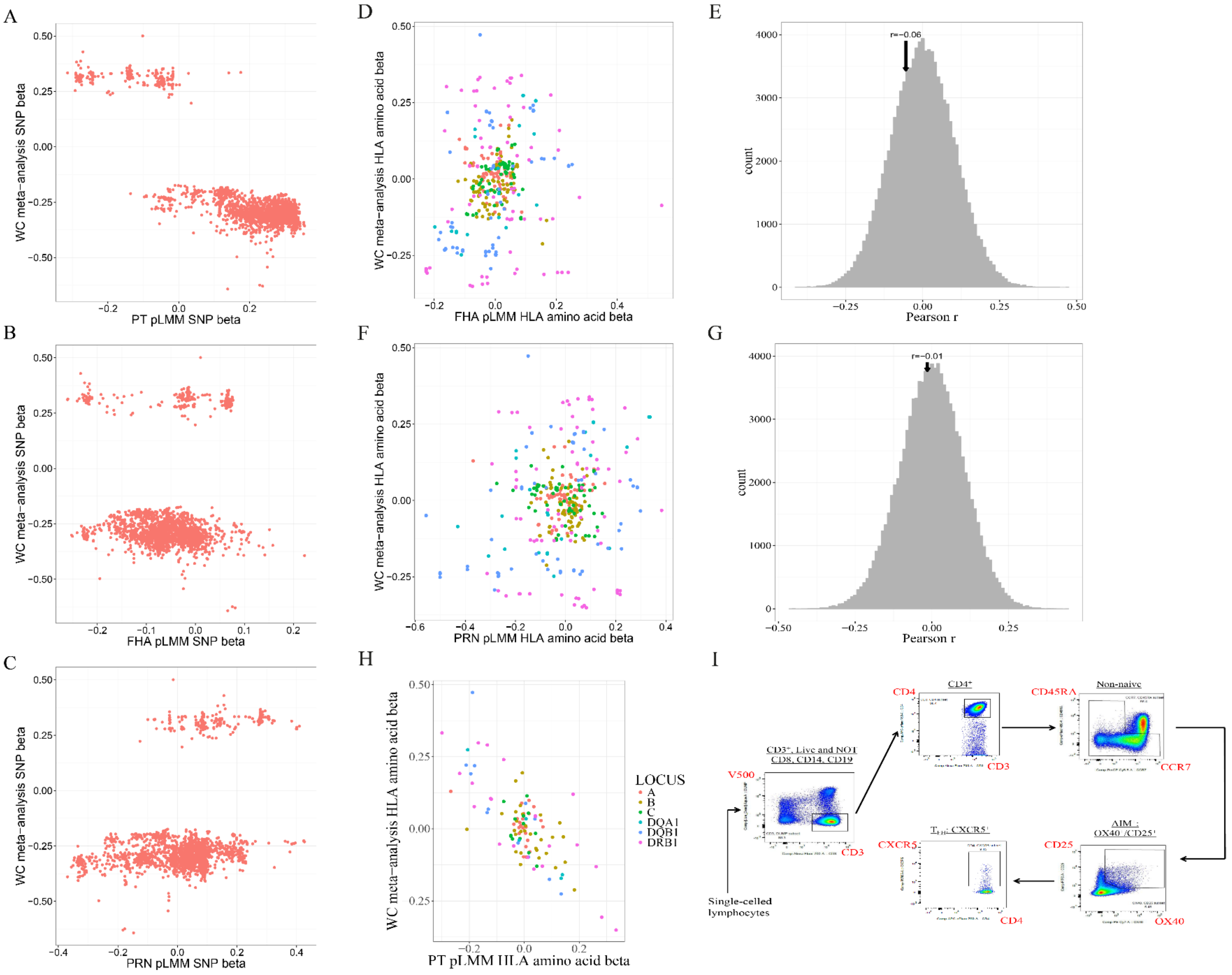
A
B
C
D
E
F
G
H
I
WC meta-analysis SNP beta
PT pLMM SNP beta
FHA pLMM SNP beta
PRN pLMM SNP beta
WC meta-analysis HLA amino acid beta
FHA pLMM HLA amino acid beta
PRN pLMM HLA amino acid beta
PT pLMM HLA amino acid beta
count
Pearson r
r=-0.06
r=-0.01
LOCUS
A
B
C
DQA1
DQB1
DRB1
Single-celled lymphocytes
CD3+, Live and NOT CD8, CD14, CD19
V500
CD3
CD4+
CD4
Non-naive
CD45RA
CCR7
AIM : OX40+/CD25+
CD25
OX40
CXCR5

**Fig. S3.**

**Exploring the functional consequence of HLA variation on response to vaccination. (A- C)** Correlation of SNP beta effect estimates between whooping cough GWAS and GWAS of PT **(A)**, FHA **(B)** and PRN **(C)** responses. Estimates were not available for pertussis GWAS for SNPs with $P > 1x10^{-5}$. Pearson's r coefficients calculated at -0.86 **(A)**, 0.19 **(B)** and 0.38 **(C)**. Individuals recruited with vaccine-related data and samples available for analysis as part of VaccGene in each cohort. **(D-G)** The beta effect estimates for association between HLA amino acid residues and FHA **(D)** and PRN **(F)** antibody responses in *VaccGene* infants are plotted against the equivalent estimates from a pertussis GWAS. Residues are colored by HLA gene. The distributions of measured Pearson r following 100,000 permutations to measure the significance of correlation between effect estimates of HLA amino acids pruned by LD ($r^2$<0.35) comparing responses against FHA **(E)** and PRN **(G)** are also shown.. **(H)** The beta effect estimates for association between HLA amino acid residues and PT antibody response in the *VaccGene* infants are plotted against the equivalent estimates from a whooping cough GWAS following pruning of the residues by LD ($r^2$<0.35). Residues are colored by HLA gene. **(I)** Gating strategy to identify AIM non-naïve $CD4^+$ T-cells and the $T_{FH}$ ($CXCR5^+$) subset. **(J-K)** Distribution of log likelihood ratios of selection with biallelic exonic and non-exonic variants in HLA-DRB1 **(J)** and HLA-C **(K)** with linkage disequilibrium estimates shown with the most exonic variant with the most evidence of selection.

**Table S1.**

Descriptive characteristics of the individuals recruited into the study from three African *VaccGene* populations.

| | Uganda | South Africa | Burkina Faso |
|---|---|---|---|
| Final genotyped number | 1,391 | 755 | 353 |
| Males, number (%) | 706 (50.8) | 384 (50.1) | 177 (50.1) |
| HIV exposed, number (%) | 130 (9.3) | 431 (57.1) | NA |
| HIV infected, number (%) | 19 (1.3) | 1 (0.1) | NA |
| Age at serum/plasma sampling, mean (s.d.*) | 54.1 (3.9) | 26.4 (3.7) | 87.4 (23.4) |
| Twin / triplet sets, number (%) | 15 (1.1) | 25 (3.3) | 0 (0) |
| **Maternal Ethnicity** | | | |
| Baganda, number (%) | 707 (50.8) | | |
| Banyankole, number (%) | 131 (9.4) | | |
| Bunyarwanda, number (%) | 76 (5.5) | | |
| Busoga, number (%) | 40 (2.8) | | |
| Batooro, number (%) | 65 (4.7) | | |
| Luo, number (%) | 78 (5.6) | | |
| Other Ugandan, number (%) | 294 (21.1) | | |
| Zulu, number (%) | | 278 (36.8) | |
| Sotho, number (%) | | 186 (24.6) | |
| Xhosa, number (%) | | 112 (14.8) | |
| Tsonga, number (%) | | 54 (7.2) | |
| Tswana, number (%) | | 51 (6.8) | |
| Venda, number (%) | | 22 (1.6) | |
| Other South African, number (%) | | 72 (5.2) | |
| Gouin, number (%) | | | 167 (47.3) |
| Karaboro, number (%) | | | 65 (18.4) |
| Turka, number (%) | | | 19 (5.4) |
| Peulh, number (%) | | | 6 (1.7) |
| Other Burkinabe, number (%) | | | 8 (2.3) |
| Unknown, number (%) | 0 | 2 (0.3) | 89 (25.2) |

*: s.d.: standard deviation
NA: not available

**Table S2.**

Summary of genotype QC steps for each population. The number of individuals and variants removed within each population are presented.

| | | Uganda | South Africa | Burkina Faso |
|---|---|---|---|---|
| Pre-QC | Total number sent for genotyping | 1,416 | 784 | 360 |
| | Number of variants typed and mapping to Build 37 | 2,328,340 | 2,328,340 | 2,328,340 |
| Individual and autosomal variant QC | Individuals failing call rate (<97%) | 2 | 2 | 1 |
| | Individuals with extreme heterozygosity (>3s.d.* around the mean) | 13 | 13 | 5 |
| | Individuals failing sex-check | 2 | 2 | 1 |
| | Individuals IBD>0.9 and not twins / triplets | 8 | 12 | 0 |
| | Autosomal variants with call rate <97% | 73,540 | 66,464 | 79,400 |
| | Autosomal variants in Hardy-Weinberg disequilibrium ($p<10^{-8}$) | 27,002 | 19,629 | 11,647 |
| Chromosome X QC | Chromosome X variants with call rate <97% | 1,552 | 1,244 | 1,599 |
| | Chromosome X variants in Hardy-Weinberg disequilibrium ($p<10^{-8}$) | 45 | 29 | 8 |
| Post-QC | Total individuals remaining | 1,391 | 755 | 353 |
| | Total autosomal variants remaining | 2,176,695 | 2,191,144 | 2,186,190 |
| | Total chromosome X variants remaining | 45,725 | 47,292 | 46,868 |

*: s.d.: standard deviations

**Table S3.**

Number of variants imputed into individual datasets divided between autosomes and the X-chromosomes. The total number of imputed variants includes all those listed in the output files direct from IMPUTE2. High quality SNPs are those only with an info score greater than 0.3 and those tested for association were only those of high quality with a minor allele frequency (MAF) greater than 0.01.

| **Autosomal variants post-imputation** | | | |
|---|---|---|---|
| Cohort | Total imputed (n) | Total high quality (n) | Total tested for association (n) |
| Uganda | 103,992,756 | 49,820,138 | 17,812,003 |
| South Africa | 103,948,407 | 39,414,539 | 20,351,192 |
| Burkina Faso | 103,543,613 | 31,941,883 | 18,537,816 |
| **X-chromosome variants post-imputation** | | | |
| Uganda | 4,257,294 | 1,792,278 | 724,497 |
| South Africa | 4,257,257 | 1,354,576 | 757,584 |
| Burkina Faso | 4,257,217 | 1,140,538 | 682,994 |

**Table S4.**

DNA samples for Maasai population.

| ID | Sex | ID | Sex | ID | Sex | ID | Sex |
|---|---|---|---|---|---|---|---|
| NA21739 | Male | NA21435 | Male | NA21582 | Male | NA21457 | Female |
| NA21775 | Female | NA21410 | Male | NA21409 | Female | NA21420 | Male |
| NA21307 | Male | NA21417 | Male | NA21476 | Female | NA21526 | Female |
| NA21524 | Female | NA21458 | Male | NA21344 | Male | NA21379 | Female |
| NA21451 | Female | NA21573 | Male | NA21357 | Female | NA21513 | Female |
| NA21367 | Male | NA21575 | Male | NA21719 | Male | NA21399 | Male |
| NA21522 | Male | NA21523 | Male | NA21770 | Female | NA21520 | Male |
| NA21693 | Female | NA21825 | Female | NA21741 | Male | NA21449 | Female |
| NA21387 | Male | NA21338 | Male | NA21596 | Male | NA21479 | Female |
| NA21740 | Male | NA21359 | Male | NA21486 | Female | NA21362 | Female |
| NA21297 | Female | NA21440 | Male | NA21448 | Male | NA21306 | Female |
| NA21743 | Male | NA21613 | Female | NA21774 | Female | NA21391 | Female |
| NA21478 | Male | NA21351 | Male | NA21650 | Female | NA21519 | Male |
| NA21722 | Male | NA21576 | Female | NA21356 | Male | NA21308 | Female |
| NA21365 | Female | NA21402 | Male | NA21632 | Female | NA21631 | Male |
| NA21577 | Male | NA21493 | Female | NA21510 | Female | NA21615 | Female |
| NA21447 | Male | NA21617 | Female | NA21583 | Male | NA21423 | Male |
| NA21768 | Female | NA21295 | Male | NA21683 | Female | NA21441 | Female |
| NA21485 | Male | NA21614 | Male | NA21769 | Female | NA21418 | Female |
| NA21333 | Female | NA21300 | Female | NA21385 | Female | NA21600 | Female |
| NA21341 | Male | NA21360 | Female | NA21584 | Male | NA21339 | Female |
| NA21352 | Male | NA21786 | Female | NA21415 | Female | NA21611 | Female |
| NA21622 | Female | NA21301 | Male | NA21616 | Male | NA21685 | Male |
| NA21679 | Female | NA21403 | Female | NA21408 | Male | NA21635 | Female |
| NA21574 | Female | NA21529 | Female | NA21298 | Male | NA21436 | Female |
| NA21744 | Male | NA21438 | Female | NA21456 | Male | NA21742 | Male |
| NA21733 | Female | NA21512 | Male | NA21580 | Female | NA21620 | Female |
| NA21424 | Female | NA21488 | Male | NA21355 | Male | NA21587 | Male |
| NA21826 | Female | NA21784 | Female | NA21473 | Female | NA21382 | Female |
| NA21686 | Female | NA21647 | Male | NA21716 | Male | NA21364 | Female |
| NA21304 | Male | NA21785 | Female | NA21717 | Female | NA21390 | Male |
| NA21634 | Male | NA21454 | Female | NA21517 | Female | NA21405 | Male |
| NA21491 | Female | NA21776 | Female | NA21335 | Male | NA21738 | Male |
| NA21368 | Female | NA21578 | Female | NA21678 | Male | NA21400 | Female |
| NA21509 | Male | NA21619 | Male | NA21443 | Male | NA21521 | Male |
| NA21599 | Male | NA21414 | Male | NA21689 | Male | NA21421 | Female |
| NA21388 | Female | NA21528 | Male | NA21723 | Female | NA21782 | Female |
| NA21318 | Male | NA21363 | Female | NA21303 | Female | NA21597 | Female |
| NA21381 | Male | NA21320 | Female | NA21489 | Female | NA21378 | Male |
| NA21649 | Male | NA21732 | Male | NA21453 | Male | NA21371 | Female |
| NA21515 | Male | NA21737 | Male | NA21336 | Female | | |
| NA21316 | Male | NA21353 | Female | NA21450 | Male | | |

**Table S5.**

Numbers of individuals with intersecting genotype, MiSeq-based 6-digit ‘G’ resolution, PacBio (potentially 8-digit resolution), or Sanger-sequence based calls.

| | | | PacBio† | | | | | | | | |
|---|---|---|---|---|---|---|---|---|---|---|---|
| | **Genotype*** | **MiSeq†** | **A** | **B** | **C** | **DRB1** | **DPB1** | **DQB1** | **DPA1** | **DQA1** | **Sanger** |
| Uganda | 330 | 330 | 0 | 0 | 0 | 0 | 0 | 0 | 0 | 0 | 47 |
| Burkina Faso | 167 | 167 | 0 | 0 | 0 | 0 | 0 | 0 | 0 | 0 | 0 |
| South Africa | 335 | 400 | 189 | 197 | 196 | 151 | 98 | 195 | 133 | 177 | 0 |
| ACB | 77 | 79 | 61 | 74 | 57 | 27 | 30 | 31 | 8 | 9 | 0 |
| GWD | 112 | 112 | 80 | 92 | 84 | 60 | 32 | 64 | 3 | 4 | 0 |
| ESN | 99 | 99 | 74 | 86 | 64 | 30 | 62 | 58 | 9 | 13 | 0 |
| MSL | 84 | 84 | 59 | 62 | 64 | 49 | 13 | 58 | 11 | 11 | 0 |
| YRI | 108 | 110 | 69 | 76 | 88 | 58 | 5 | 85 | 6 | 7 | 0 |
| LWK | 97 | 97 | 77 | 64 | 82 | 52 | 2 | 81 | 4 | 5 | 0 |
| ASW | 54 | 62 | 45 | 43 | 50 | 27 | 0 | 50 | 0 | 0 | 0 |
| MKK | 134 | 166 | 141 | 142 | 87 | 54 | 105 | 132 | 14 | 20 | 0 |
| **Total** | **1597** | **1706** | **795** | **836** | **772** | **508** | **347** | **754** | **188** | **246** | **47** |

*: Genotype data was either available from Omni2.5M genotype calling or next generation sequence data available intersecting with HLA type data of any type.

†: All PacBio data was available on individuals who also had MiSeq data, but not necessarily genotype data.

**Table S6.**

Novel protein coding alleles discovered in the combined 1000Gp3-*VaccGene* dataset. Genbank accession numbers try to represent a single sequence for each novel allele submission.

| Gene | Reported Allele | Genbank Accession | WHO Reference | New Allele |
|---|---|---|---|---|
| | 02:XX | MH973915 | | NA* |
| | 02:XX | MH973917 | | NA* |
| | 02:XX | MH973918 | | NA* |
| HLA-A | 23:XX | KU668724 | 10031574 | 23:73 |
| | 26:XX | KU668725 | 10031563 | 26:121 |
| | 30:XX | MK032383 | | NA |
| | 32:XX | MG429694 | | 32:106 |
| HLA-B | 15:XX | In submission | | NA |
| | 15:10:XX | MF170525 | | NA |
| | 02:XX | In submission | | NA |
| | 02:10:XX | MH544312 | | 02:10:04 |
| HLA-C | 04:XX | MH544322 | 10039321 | 04:368 |
| | 07:43P | KX017417 | 10032163 | 07:43:02 |
| | 07:XX | MG769797 | | 07:629 |
| | 01:XX | In submission | | NA* |
| | 01:XX | In submission | | NA* |
| | 01:XX | MF170464 | | 01:15 |
| | 02:XX | MF170461 | | NA* |
| | 02:XX | MF170462 | | 02:09 |
| | 02:XX | MF170456 | | NA* |
| | 02:XX | MF170459 | | NA* |
| HLA-DPA1 | 02:XX | MH536331 | | 02:12 |
| | 02:07:XX | NA | | 02:07:01* |
| | 03:XX | In submission | | NA* |
| | 03:XX | MF170453 | | NA* |
| | 03:01:XX | NA | | 03:01:02* |
| | 03:02P | MF170463 | | NA |
| | 04:XX | NA | | 04:02 |
| | 02:01:XX | MH974010 | | NA |
| | 03:XX | In submission | | NA |
| | 11:XX | NA | | 654:01 |
| | 55:01:XX | MG805509 | | 55:01:02 |
| HLA-DPB1 | 104:01:XX | In submission | | NA |
| | XX | KU668852 | 10031694 | 558:01 |
| | XX | KU668853 | 10031698 | 561:01 |
| | XX | NA | | 584:01 |
| | 01:XX | MK442249 | 10043295 | NA |
| | 01:XX | MF170427 | | NA |
| | 01:01:XX | In submission | | |
| HLA-DQA1 | 04:XX | MK442251 | | NA |
| | 04:XX | MF170425 | | NA |
| | 04:XX | MF170426 | | NA |
| | 04:XX | In submission | | NA |
| | 02:XX | KU668797 | 10031657 | 02:70* |
| | 02:XX | | | 02:70* |
| | 02:01P | MK058598 | | NA |
| HLA-DQB1 | 04:XX | MH536304 | | 04:52 |
| | 04:XX | In submission | | NA |
| | 04:02:XX | NA | | 04:02:13 |

| | | | | |
|---|---|---|---|---|
| HLA-DRB1 | 03:XX | KU668802 | 10031673 | 03:131 |
| | 10:01:XX | MK192150 | 10042610 | NA |
| HLA-DRB3 | 01:XX | MK058635 | | NA |

*: These alleles may represent the same novel allele as others listed in the class but sequence reads are in the process of independent evaluation

### Table S7.

Results from manual and automated step-wise modelling of class II HLA variants with five vaccine responses including principal components and time between sampling and vaccination as covariates. The reported SNP variants all had info scores greater than 0.8 (rs73727916 info 0.83-0.91 across the three cohorts; rs147857322 0.97-0.99 and rs34951355 0.88-0.91).

| | Method | Variant 1[*] | Variant 2[*] | | Variant 3[*] | Variant 4[*] | $P_{LMM}$ | $P_{uni}$[†] | $P_{multi}$[‡] | BIC[§] |
|---|---|---|---|---|---|---|---|---|---|---|
| PT | Manual for HLA-DRB1 and HLA-DRB3 | rs73727916 | - | | - | - | 3.6x10$^{-26}$ | 8.1x10$^{-30}$ | - | 6442.55 |
| | | DRB3-74Gln↾ | - | | - | - | 4.2x10$^{-25}$ | 2.0x10$^{-28}$ | - | 6453.00 |
| | | HLA-DRB3*02:02:01G↾ | HLA-DRB3*03:01:01G↾ | | - | - | - | - | 2.0x10$^{-28}$ | 6451.49 |
| | | DRB1-233Thr | - | | - | - | - | 1.7x10$^{-26}$ | - | 6457.75 |
| | **Final** | **rs73727916** | **DRB3-74Gln** | | - | - | - | - | 3.3x10$^{-35}$ | **6420.17** |
| FHA | Manual for HLA-DRB1 | HLA-DRB1*08:04:01↾ | - | | - | - | 4.8x10$^{-15}$ | 5.7x10$^{-16}$ | - | 6492.47 |
| | | DRB1-74Leu↾ | - | | - | - | 4.9x10$^{-15}$ | 7.3x10$^{-16}$ | - | 6492.95 |
| | Automated | HLA-DRB1*15:03:01G | - | | - | - | 2.6x10$^{-8}$ | 6.3x10$^{-10}$ | - | 6519.79 |
| | **Final** | **HLA-DRB1*08:04:01** | HLA-DRB1*15:03:01G | | - | - | - | - | 1.8x10$^{-21}$ | **6470.25** |
| PRN | Manual for HLA-DQB1 | rs147857322 | - | | - | - | 4.2x10$^{-23}$ | 1.1x10$^{-25}$ | - | 4469.76 |
| | | DQB1-74Ser↾ | - | | - | - | 1.8x10$^{-21}$ | 4.6x10$^{-25}$ | - | 4472.52 |
| | | DQB1*05:01:01G↾ | HLA-DQB1*04:02:01↾ | | - | - | - | - | 3.7x10$^{-27}$ | 4463.84 |
| | Automated | HLA-DRB1*11:02:01 | - | | - | - | - | 2.3x10$^{-13}$ | - | 4525.74 |
| | | DQB1-175Glu | - | | - | - | - | 7.4x10$^{-20}$ | - | 4596.24 |
| | **Final** | **rs147857322** | **DQB1-74Ser** | | **HLA-DRB1*11:02:01** | **DQB1-175Glu** | - | - | 1.4x10$^{-38}$ | **4415.14** |

[*]**:** Variants associated with each trait through identified through the pLMM or univariate modelling approaches

[†]**:** The univariate model tested only on individuals restricted by IBD (< 0.2)

[‡]**:** Where multiple variants are tested in a conditional multivariate model the omnibus p-value is shown ($p_{multi}$)

[§]: The Bayesian Information Criterion was calculated for all models. The most parsimonious model will have a BIC closer to 0. The model with the lowest BIC for each phenotype at each locus is shown in red and the BIC of the final model is shown in bold.

↾ For each locus and each trait any associated HLA amino acid was also tested in a univariate or multivariate model as described by classical HLA alleles that contain that residue. Although improved in the model when contained alone, DQB1-74Ser improved the final model when including the SNP variant in PRN.

**Table S7.** continued

| | | | | | | | | | |
|---|---|---|---|---|---|---|---|---|---|
| DT | Manual for HLA-DRB1 | rs34951355 | - | - | - | $1.5x10^{-26}$ | $1.2x10^{-30}$ | - | 6347.03 |
| | **Final** | **rs34951355** | - | - | - | $1.5x10^{-26}$ | $1.2x10^{-30}$ | | **6347.03** |
| HBsAg | Manual for HLA-DRB1 | DRB1-74Arg† | - | - | - | $6.3x10^{-14}$ | $1.9x10^{-18}$ | - | 5276.06 |
| | | HLA-DRB1*03:02:01 | - | - | - | - | $6.3x10^{-15}$ | - | 5292.09 |
| | | HLA-DRB1*03† | - | - | - | - | $1.9x10^{-18}$ | - | 5276.06 |
| | | HLA-DRB1*03:02:01† | HLA-DRB1*03:01:01G† | - | - | - | - | $3.1x10^{-18}$ | 5279.81 |
| | Manual for HLA-DPB1 | DPB1-85Gly† | - | - | - | $1.2x10^{-10}$ | $6.9x10^{-13}$ | - | 5301.34 |
| | | HLA-DPB1*105:01 | - | - | - | - | $2.9x10^{-6}$ | - | 5331.00 |
| | | HLA-DPB1*02:01:02† | HLA-DPB1*04:01:01G† | HLA-DPB1*18:01† | - | - | - | $4.3x10^{-9}$ | 5318.41 |
| | Automated | DRB1-67Phe† | - | - | - | - | $1.1x10^{-11}$ | - | 5306.81 |
| | | HLA-DRB1*08:04:01† | HLA-DRB1*09:01:02G† | HLA-DRB1*11:01:02† | - | - | - | $1.8x10^{-11}$ | 5307.77 |
| | | DPB1-35Tyr | - | - | - | - | $1.7x10^{-13}$ | - | 5298.59 |
| | **Final** | **DRB1-74Arg** | **DPB1-85Gly** | **DRB1-67Phe** | **DPB1-35Tyr** | - | - | $6.9x10^{-32}$ | **5219.97** |

§: The Bayesian Information Criterion was calculated for all models. The most parsimonious model will have a BIC closer to 0. The model with the lowest BIC for each phenotype at each locus is shown in red and the BIC of the final model is shown in bold.

† For each locus and each trait any associated HLA amino acid was also tested in a univariate or multivariate model as described by classical HLA alleles that contain that residue.

**Table S8.**

Signals of association between class II HLA variants and three vaccine response traits demonstrating significant evidence of heterogeneity.

| | | Uganda | | | South Africa | | | Burkina Faso | | | | | |
|---|---|---|---|---|---|---|---|---|---|---|---|---|---|
| | **Variant** | **Beta** | **s.e.** | **MAF*** | **Beta** | **s.e.** | **MAF*** | **Beta** | **s.e.** | **MAF*** | $P_{FE}$† | $P_{RE}$‡ | $P_{Q}$§ |
| **PT** | DRB3-74Gln | -0.44 | 0.04 | 0.48 | -0.04 | 0.05 | 0.40 | -0.34 | 0.08 | 0.35 | $3.1x10^{-28}$ | 0.05 | $1.3x10^{-9}$ |
| | rs73727916 | 0.43 | 0.04 | 0.40 | 0.16 | 0.06 | 0.37 | 0.24 | 0.08 | 0.52 | $6.1x10^{-27}$ | $3.1x10^{-3}$ | $1.8x10^{-4}$ |
| **FHA** | HLA-DRB1*08:04:01 | 0.49 | 0.09 | 0.05 | 0.23 | 0.14 | 0.04 | 0.93 | 0.13 | 0.08 | $1.2x10^{-16}$ | $2.2x10^{-3}$ | $1.1x10^{-3}$ |
| **DT** | rs34951355 | 0.73 | 0.07 | 0.08 | 0.41 | 0.08 | 0.12 | 0.29 | 0.11 | 0.12 | $1.2x10^{-29}$ | $3.5x10^{-4}$ | $3.9x10^{-4}$ |

* minor allele frequency
† p-value calculated from a fixed effects meta-analysis combining the effects across the three studied populations
‡ p-value calculated from a random effects meta-analysis combining the effects across the three studied populations
§ p-value of evidence of heterogeneity between the three studied population calculated from the Cochran's Q statistic

**Table S9.**

Characteristics of donors and PBMC samples used for PT-specific $T_{FH}$ assay.

| | **DRB1-233Arg (Number (%))** | **DRB1-233Thr (Number (%))** |
|---|---|---|
| Final number | 15 | 14 |
| Males | 7 (46.7) | 6 (42.9) |
| Mean age at sampling in years | 46.1 (23.7)† | 43.2 (24.3) † |
| Mean storage time of PBMCs in years | 3.87 (2.1) † | 3.81 (2.7) † |
| **<u>Participant Ethnicity</u>** | | |
| Asian | 0 | 1 (7.1) |
| Pacific Islander | 1 (6.7) | 3 (21.4) |
| Black | 6 (40.0) | 0 |
| Hispanic / Latino | 3 (20.0) | 0 |
| White | 4 (26.7) | 9 (64.2) |
| Unknown | 1 (6.7) | 1 (7.1) |
| **4-Digit HLA-DRB1 Alleles** | | |
| 01:01 | 0 | 9 (32.1) |
| 01:02 | 0 | 2 (7.1) |
| 03:01 | 4 (13.3) | 0 |
| 03:02 | 1 (3.3) | 0 |
| 03:17 | 1 (3.3) | 0 |
| 11:01 | 7 (23.3) | 0 |
| 11:02 | 4 (13.3) | 0 |
| 11:04 | 4 (13.3) | 0 |
| 13:01 | 4 (13.3) | 0 |
| 13:02 | 1 (3.3) | 0 |
| 13:03 | 2 (6.7) | 0 |
| 13:04 | 2 (6.7) | 0 |
| 15:01 | 0 | 9 (32.1) |
| 15:02 | 0 | 7 (25.0) |
| 15:09 | 0 | 1 (3.6) |

†: values given in mean (standard deviation)

**Table S10.**

Breadth of PT-peptides binding to associated HLA-DRB1 alleles compared to the breadth of peptides derived from TT binding to the same alleles.

| | Allele | PT Breadth† | TT Breadth† | *P*-value |
|---|---|---|---|---|
| **DRB1-233Thr** | DRB1*01:01 | 3.1 | 4.6 | |
| | DRB1*01:02 | 24.2 | 26.3 | |
| | DRB1*15:01 | 3.5 | 11.3 | |
| | DRB1*15:03 | 3.1 | 8.1 | |
| | Average | 8.5 (+/- 10.5) | 12.6 (+/- 9.5) | NS |
| **DRB1-233Arg** | DRB1*03:01 | 22.03 | 26.7 | |
| | DRB1*11:01 | 9.3 | 6.5 | |
| | DRB1*11:02 | 0.4 | 7.5 | |
| | DRB1*13:02 | 12.3 | 22.4 | |
| | Average | 11.0 (+/- 8.9) | 15.8 (+/- 10.3) | NS |

†:Values represent % of peptides derived from the corresponding toxin that are predicted to bind with high affinity (+/- standard deviation) calculated according to the 5th percentile rank in IEDB.

**Additional Data Table 1 (Separate File)**

Association statistics from FE meta-analyses for all extended MHC variants (bi-allelic SNPs, HLA alleles and amino acids) in the three African populations for the five vaccine antibody responses with GWAS significant associations. SNPs are coded in build 37 coordinates as 'chromosome':'base pair':'minor allele':'major allele'. Amino acids are coded as 'Gene'_'AA1'_'full length position'_'amino acid present'_'coding sequence position'. Coding sequence position was used in the main text. HLA alleles are coded as 'Gene'_'6 digit G Allele'.

**Additional Data Table 2 (Separate File)**

Allele-specific statistics comparing imputed HLA allele calls from HLA*IMP:02 to sequence-based 6-digit ‘G’ typing divided by population. Calls are compared at 4-digit level of resolution and presented in 4-digit format. Locus A and allele 0101 refers to HLA-A*01:01 for example. New alleles defined through HLA typing are donated as XX and are detailed in **Table S6**.

**Additional Data Table 3 (Separate File)**

Allele-specific statistics comparing imputed HLA allele calls from HLA*IMP:02G to sequence-based 6-digit 'G' typing divided by *VaccGene* population. Calls are compared at 4-digit level of resolution and presented in 4-digit format for comparison to Additional Data Table 2. Locus A and allele 0101 refers to HLA-A*01:01 for example. New alleles defined through HLA typing are donated as XX and are detailed in **Table S6**.

**Additional Data Table 4 (Separate File)**

Allele-specific statistics comparing imputed HLA allele calls from HLA*IMP:02G to imputed HLA allele calls from the Broad Multi-Ethnic reference panel divided by *VaccGene* population. Calls are compared at 4-digit level of resolution and presented in 4-digit format for comparison to Additional Data Table 2. Locus A and allele 0101 refers to HLA-A*01:01 for example. New alleles defined through HLA typing are donated as XX and are detailed in **Table S6**.

**Additional Data Table 5 (Separate File)**

Summary beta, standard error and *P*-values for fixed effects meta-analysis of *cis*-QTL analyses for each of eight major class I and II *HLA* genes
